# Progression GWAS followed by functional characterization implicates E3 ubiquitin ligase *TRIM2* as a potential genetic modifier of Parkinson’s disease progression

**DOI:** 10.1101/2025.02.21.25322301

**Authors:** Ameya S. Kulkarni, Dan Apicco, Saleh Tamim, Lauren Gibilisco, Layan Nahlawi, Rachel Lucia, Sujana Ghosh, Samantha Lent, Yanyu Liang, Saurabh Khasnavis, Laura Smith, Joshua Stender, Priyanka Vijay, Cindy Zadikoff, Justin Wade Davis, Jan Stoehr, Hyun Ji Noh

## Abstract

Parkinson’s Disease (PD) is a progressive neurodegenerative disorder, affecting 10 million people worldwide. While genome-wide association studies (GWAS) have identified many genetic variants associated with PD incidence, the genetics underlying PD progression are poorly understood. Here, we aim to address this gap by performing GWAS on longitudinal clinical metrics from well-defined PD cohorts. Specifically, we identify 8 novel GWAS genes for PD progression, including *TRIM2,* which encodes an E3 ubiquitin ligase with loss-of-function mutations that cause neuropathy. Functional genomics data suggest that the GWAS SNPs in the locus of *TRIM2* regulate its expression across several PD-relevant brain regions. Further, we show that *TRIM2* knockdown and overexpression in primary neurons regulate neurofilament light (NF-L) levels and α-synuclein aggregation-the primary neuropathological hallmark of PD. Peripheral proteomic analysis of a genetically defined PD patient cohort demonstrates increased NF-L protein levels in the plasma and cerebrospinal fluid of *TRIM2* SNP carriers, corroborating the role of TRIM2 in NF-L regulation. Overall, by integrating PD progression GWAS with transcriptomic, eQTL analyses, and functional data in PD cellular models, we identify new targets including *TRIM2* that may influence the progression rate of PD.

## INTRODUCTION

Parkinson’s disease (PD) is a progressive neurodegenerative disorder that impairs a range of motor and non-motor functions. While it is predicted to impact 14 million patients worldwide by 2040, current therapies treat the resulting symptoms but do not modify disease progression^1^. Understanding the pathological mechanisms underlying PD progression may discover new targets and therefore accelerate the development of ‘disease-modifying’ therapies.

Large-scale genome-wide association studies (GWAS) have identified at least 90 genetic loci associated with PD, suggesting hundreds of potential risk genes contributing to PD incidence^2^. While these genes reveal impaired molecular mechanisms that might lead to PD, such risk loci do not necessarily imply a causal relationship, nor confirm that these genes continue to be involved in the disease progression once initiated. Recent evidence suggests that genetic risk factors and progression factors may be distinct, as there was minimal overlap in gene candidates from GWAS risk and progression studies for Alzheimer’s disease^3^.

Currently, little is known about the genetics underlying PD progression, with only a few genes previously reported^4–7^. In contrast to conventional case-control GWAS, progression GWAS can reveal genes and processes that participate longitudinally and actively shape pathobiological progression of disease. However, the unavailability of large-scale longitudinal clinical and genetic data from well-defined patient cohorts has been a major challenge to the identification of genetic modifiers of PD progression.

The recent release of PD cohorts with longitudinal clinical measurements is enabling in-depth studies of disease progression. These cohorts include expert-rated metrics such as Movement Disorder Society-Unified Parkinson’s Disease Rating Scale (MDS-UPDRS) paired with omics datasets. The Parkinson’s Progression Markers Initiative (PPMI) provides a multi-omics dataset linked with deep clinical phenotyping, neuroimaging, and biochemical measurements over several years. PPMI also includes ‘on-state’ and ‘off-state’ MDS-UPDRS scores to track more accurate clinical decline in the absence of symptomatic medication masking the true severity of clinical symptoms. Accelerating Medicines Partnership-Parkinson’s Disease (AMP-PD) is another clinical data initiative that provides multi-omics datasets from multiple independent PD cohorts, harmonizing key clinical measurements.

Advances in animal and cellular models of PD are also increasing our ability to study the molecular mechanisms associated with disease progression^8^. In the brain of PD patients, alpha-synuclein (α-syn), a protein that normally regulates neurotransmitter vesicle release at presynaptic terminals, becomes aberrantly misfolded and accumulates in large, lipid-rich intraneuronal aggregates, which are also referred to as Lewy bodies (LBs) and Lewy neurites. The spatiotemporal accumulation and spreading of α-syn pathology correlates with both neuronal loss and clinical progression in PD, suggesting that LB formation is a central driver of PD progression^9,10^. The process of progressive α-syn pathology formation can be recapitulated *in vivo* by injection of recombinant preformed fibrils (PFFs) of α-syn into mouse brain tissue, inducing a progressive spread of α-syn aggregates reminiscent of Lewy bodies, accompanied by motor symptoms and neuronal loss^11,12^. Similarly, PFF treatment also induces α-syn aggregation in primary neurons in vitro, providing a cellular model for characterizing candidate genes that might influence the formation of α-syn aggregates and therefore may show potential for therapeutic disease modification^13–15^.

Here, we present two longitudinal GWAS (AMP-PD and PPMI) together with re-analyses of PD cellular model transcriptomes^8^, which identify 4 candidate genetic modifiers of PD progression, including tripartite motif containing 2 (*TRIM2)*. Intersecting with PFF-treated mouse brain transcriptomes, GTEx eQTL brain data and chromosomal conformation data prioritizes *TRIM2* as a candidate genetic modifier of PD. The association of *TRIM2* locus with PD clinical phenotypes is confirmed in two additional human cohorts (Iwaki *et al*. and Genomics England). Following up on these genetic findings, we further demonstrate the role of TRIM2 in regulating neurofilament light (NF-L) accumulation and α-synuclein aggregation in cellular models of PD. Lastly, our peripheral proteomic analysis of PD patients suggests a link between *TRIM2* SNPs and NF-L abundance. Taken together, our results identify *TRIM2* as a strong candidate for genetic modifier of PD clinical progression, along with other novel candidate genes and pathways, providing new insights into the molecular mechanisms of PD.

## RESULTS

### Two longitudinal GWAS identify 8 genes significantly associated with PD progression

To identify genetic modifiers of PD progression, we conducted two distinct GWAS on the clinical data from longitudinal patient cohorts: the AMP-PD (N=1,114) and PPMI (N=319) datasets. In the AMP-PD dataset, we focused on identifying variants that are associated with the rate of change in the overall PD clinical rating over time (PD progression) in as many patients as possible, leveraging the relatively large sample size. In the PPMI dataset, which is a subset of the AMP-PD cohort that contains more detailed clinical metrics, we exclusively used the MDS-UPDRS from a drug-naïve state or ‘off-state’ (i.e., measured after anti-PD medication effects are ‘washed off’ from patients) to minimize the confounding effect of PD symptomatic medication on PD progression metrics. For both analyses, we used the total MDS-UPDRS scores measured at different time points within each patient to derive the phenotype (**Figure 1a-b**) and queried for SNPs associated with the rate of clinical progression (**Online Methods, Table 1**).

**Figure 1:**
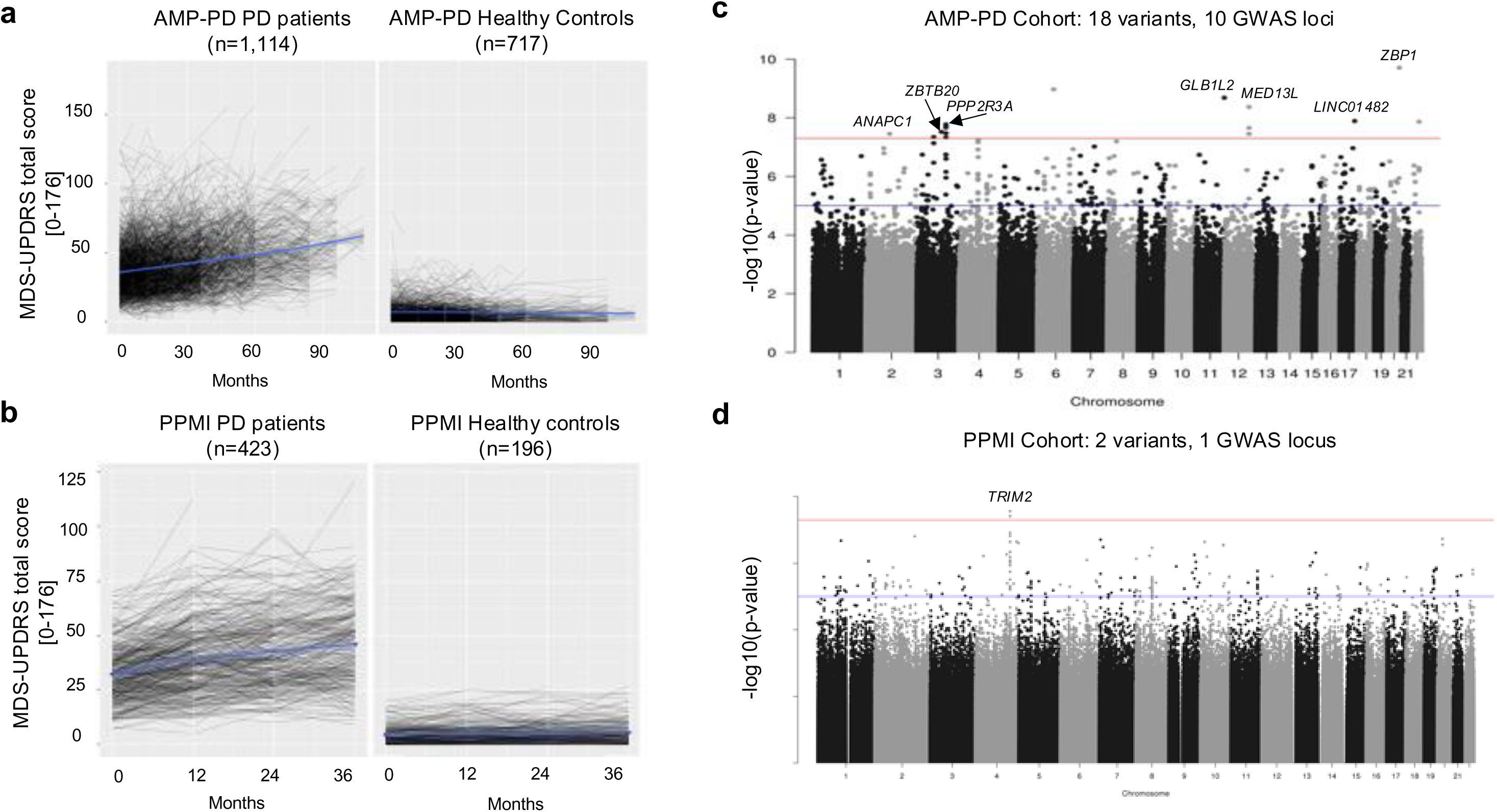
Longitudinal GWAS analyses identify genes associated with PD progression, defined as the rate of change in total MDS-UPDRS. **a-b**. In AMP-PD and PPMI datasets, Movement Disorder Society-Unified Parkinson’s Disease Rating Scale (MDS-UPDRS) total scores (sum of parts I, II, III) increase over time among PD patients, while the scores remain relatively unchanged in healthy controls. **c-d.** PD progression GWAS identify 18 variants in (c) AMP-PD and (d) two variants in PPMI that are significantly associated with PD progression.

**TABLE 1.** Significantly associated variants identified from PD progression GWAS.

| No. | Variant ID | Dataset | Allele frequency in dataset | Allele frequency in gnomAD | P-value | Beta | Annotated Genes |
| --- | --- | --- | --- | --- | --- | --- | --- |
| 1 | rs72729647 | PPMI | 0.219 | 0.190 | 2.75E-08 | 4.388 | *TRIM2* |
| 2 | rs62323742 | PPMI | 0.298 | 0.263 | 3.88E-08 | 3.897 | *TRIM2* |
| 3 | rs574099767 | AMP-PD | 0.011 | 0.010 | 3.54E-08 | 7.629 | *ANAPC1* |
| 4 | rs73107779 | AMP-PD | 0.010 | 0.007 | 4.55E-08 | 7.706 | - |
| 5 | rs142084086 | AMP-PD | 0.013 | 0.011 | 3.03E-08 | 6.852 | *ZBTB20* |
| 6 | rs116741793 | AMP-PD | 0.010 | 0.008 | 4.63E-08 | 7.701 | - |
| 7 | rs147934961 | AMP-PD | 0.009 | 0.006 | 2.15E-08 | 8.251 | - |
| 8 | rs116597875 | AMP-PD | 0.009 | 0.006 | 2.15E-08 | 8.251 | - |
| 9 | rs149169946 | AMP-PD | 0.008 | 0.007 | 1.70E-08 | 8.970 | - |
| 10 | rs149239932 | AMP-PD | 0.008 | 0.007 | 1.70E-08 | 8.970 | *PPP2R3A* |
| 11 | rs142236497 | AMP-PD | 0.008 | 0.007 | 3.31E-08 | 8.785 | - |
| 12 | rs192758048 | AMP-PD | 0.013 | 0.006 | 1.07E-09 | 7.823 | - |
| 13 | rs147142161 | AMP-PD | 0.013 | 0.010 | 2.08E-09 | 7.287 | *GLB1L2* |
| 14 | rs139365555 | AMP-PD | 0.012 | 0.008 | 4.29E-09 | 7.667 | *MED13L* |
| 15 | rs147826235 | AMP-PD | 0.013 | 0.009 | 3.54E-08 | 6.959 | - |
| 16 | rs116952358 | AMP-PD | 0.013 | 0.008 | 2.22E-08 | 6.959 | - |
| 17 | rs118168984 | AMP-PD | 0.013 | 0.009 | 3.54E-08 | 6.959 | - |
| 18 | rs143693407 | AMP-PD | 0.010 | 0.008 | 1.29E-08 | 8.219 | *LINC01482* |
| 19 | rs141673834 | AMP-PD | 0.010 | 0.006 | 1.97E-10 | 8.617 | *ZBP1* |
| 20 | rs200474818 | AMP-PD | 0.013 | 0.007 | 1.36E-08 | 7.314 | - |

### AMP-PD GWAS signals arise from rare variants

The GWAS in AMP-PD identified 18 genetic variants that are significantly associated with PD progression at the genome-wide level (P < 5E-08) (**Figure 1c**, **Table 1**, **Supplementary Table 1, Supplementary Figure 1**). These variants were relatively rare in our PD dataset (allele frequency [AF] < 0.02), and their frequencies were slightly above those reported in the population database gnomAD (**Table 1**). 13/18 variants reside within 5 distinct genomic regions, and 5/18 variants did not have linkage disequilibrium (LD) pairs in our dataset (**Supplementary Table 1**). 6/18 are intronic variants of the following protein-coding genes: mediator complex subunit 13L *(MED13L)* encoding a transcriptional coactivator for RNA polymerase II-transcribed genes, anaphase-promoting complex subunit 1 (*ANAPC1)* encoding a cell cycle-regulated E3 ubiquitin ligase, zinc finger and BTB domain containing 20 *(ZBTB20)* encoding a transcriptional repressor for neurogenesis, protein phosphatase 2 regulatory subunit B’’alpha *(PPP2R3A)* encoding a substrate selectivity and catalytic activity modulator, galactosidase beta 1 like 2 *(GLB1L2)* which is predicted to enable glycoside hydrolase activity in vacuoles, and Z-DNA-binding protein 1 *(ZBP1*) encoding a key activator of tumor necrosis factor-alpha (TNF)-induced necroptosis^16–18^. Another variant resides in the upstream of long intergenic non-protein coding RNA 1482*, LINC01482*.

### PPMI GWAS gene TRIM2 shows association signals in common and rare variants

The PPMI cohort contains more detailed phenotyping information on clinical decline than other AMP-PD datasets. Taking advantage of the refined phenotypes, we hypothesized that investigating genomic association in the PPMI dataset may identify additional genes associated with PD progression in the absence of any potential confounding effects of PD symptomatic medication.

GWAS in the PPMI cohort identified two common variants: rs72729647 (P=2.75E-8, AF=0.219) and rs62323742 (P=3.88E-8, AF=0.298) to be associated with PD progression at the genome-wide significant level (**Figure 1d**, **Table 1**, **Supplementary Table 1, Supplementary Figure 1**). The two variants are 44kb apart with moderate LD (r^2^=0.43), residing in the introns of an E3 ubiquitin ligase gene, tripartite motif containing 2 (*TRIM2*)^19^. Conditional analysis found no evidence of multiple independent signals in this locus (**Supplementary Note**).

Pathway analyses on the genes from the genomic regions with suggestive associations in the AMP-PD and the PPMI GWAS suggested potential roles of these genes in biological functions related to PD progression (**Supplementary Table 2, Supplementary Table 3, Supplementary Table 4, Supplementary Note**).

To evaluate the aggregate effect of rare, protein-coding variants, we also performed gene-based test. While 956 genes showed nominal associations with PD progression (P < 0.05, **Supplementary Table 5**), no genes were significantly associated after multiple testing correction (FDR < 0.05). However, *TRIM2* missense mutations showed an association with slower PD progression (SKAT-O P=0.008, **Supplementary Note**, **Supplementary Table 5**, **Supplementary Figure 2**), corroborating that the *TRIM2* locus may contribute to PD progression.

### Expression changes of 4 PD progression GWAS genes correlate with Lewy body formation

In combination, the PD progression GWAS of the AMP-PD and PPMI cohorts identified 8 genes as potential genetic modifiers of PD progression: *ANAPC1, GLB1L2, MED13L, PPP2R3A, ZBP1, ZBTB20, LINC01482,* and *TRIM2* (**Table 1**). To prioritize our GWAS results for functional relevance, we next hypothesized that some of these 8 PD progression GWAS genes might exhibit differential expression in response to α-syn aggregation. We re-analyzed publicly available bulk RNA-seq data from primary mouse hippocampal neurons treated with PFFs^8^. We conducted 15 pairwise comparisons of transcriptomes from neurons that underwent 0 day (PBS), 1 day (D1), 3 days (D3), 7 days (D7), 14 days (D14), and 21 days (D21) of PFF treatment, where prominent changes in α-syn aggregates reminiscent of LB formation (*e.g.* sequestration of lysosome and mitochondria, signs of neuronal death) were observed between D14 and D21 (**Supplementary Note**). 3,299 genes were significantly differentially expressed with ≥2-fold changes (PFF-induced DEGs, adjusted P < 0.05 at any steps of α-syn aggregate induction process) (**Figure 2a**, **Supplementary Table 6)**.

**Figure 2:**
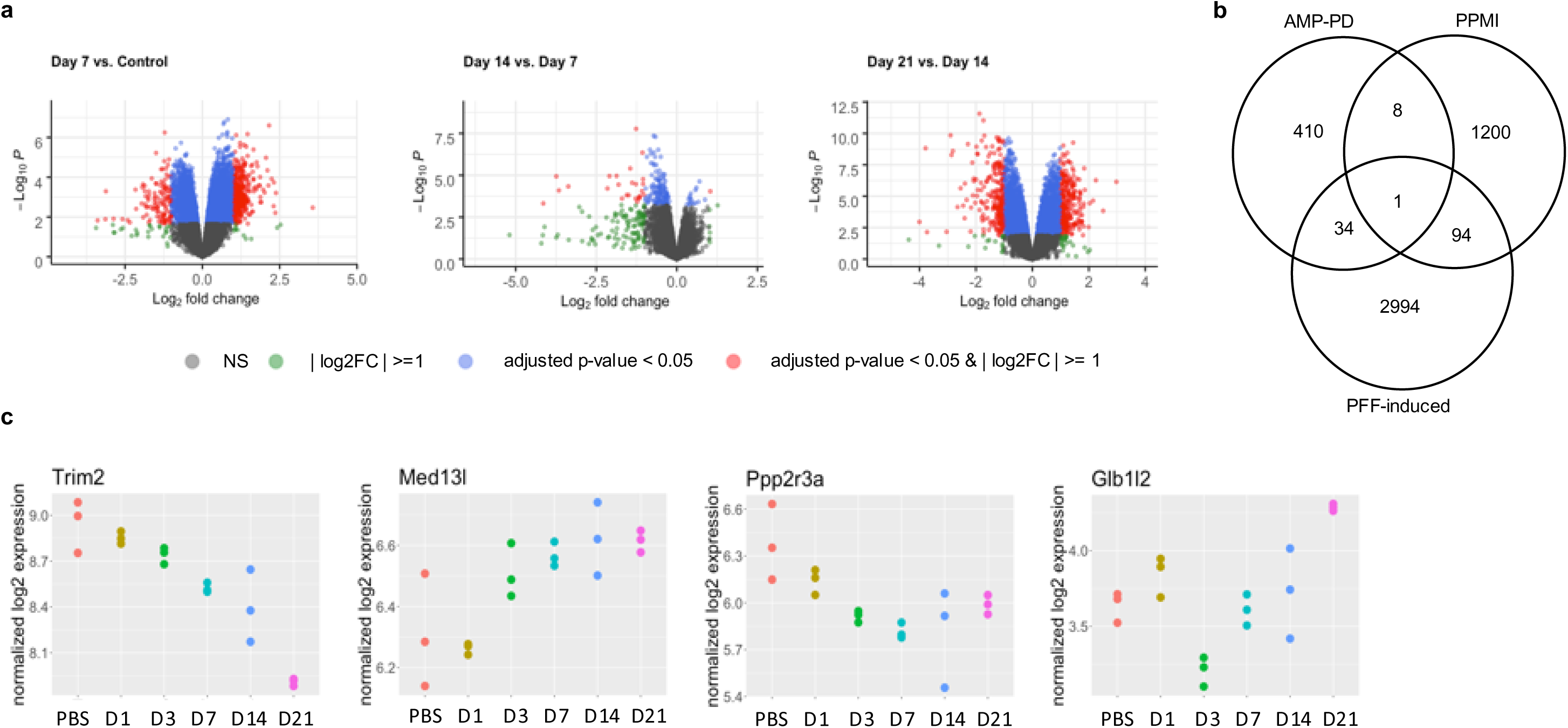
Integration of GWAS and PFF-induced transcriptome identifies four PD progression genes differentially expressed in response to PD pathology in neurons. **a.** 15 pairwise differential analyses of time-course transcriptomes in neurons that underwent 21 days of a-syn aggregation identify >3,000 PFF-induced DEGs. 3 key comparisons are shown here. PBS is baseline timepoint before PFF treatment. D: days passed after PFF-treatment. NS: Not significant. FC: fold-change. **b.** 3-way comparison of AMP-PD and PPMI genes with suggestive association (P < 1E-5) and PFF-induced DEGs identify 137 candidate genes for PD progression supported by 2+ datasets. **c.** Trim2 shows consistent decrease in expression over the course of a-syn aggregation. In 10/15 comparisons, the differential expression was significant. Med13l shows a trend of increased expression after pS129+ aggregate observation at D4, with a statistically significant increase at the timepoints (D21, D14, and D7) vs. PBS and D1 time points and at D3 vs. PBS. Ppp2r3a has decreased expression with increasing length of PFF treatment, with significant changes at D3, D7, D14, and D21 vs. PBS. Glb1l2 has decreased expression after 3 days of PFF treatment, then increased again with increasing length of PFF treatment, with significant changes at D21 vs. all other timepoints, D3 vs. D1 and PBS, and D14 vs. D3.

Pathway analyses on the 3,123 PFF-induced DEGs with human orthologs and comparison with PD progression genes of suggestive association (P <1E-5) from our GWAS detected specific pathways and genes previously associated with PD **(Supplementary Note, Supplementary Table 6, Supplementary Table 7, Figure 2b)**.

4 of the 8 candidate genes detected in our PD progression GWAS were among the top PFF-induced DEGs in PFF-treated primary neurons (**Figure 2c**). (1) Trim2 expression notably decreases as PFF treatment time increases. In 10/15 comparisons, this decrease was significant (1.2-2-fold changes). (2) Med13l expression increases, particularly after pS129+ α-syn aggregate observation at D4. (3) Ppp2r3a expression decreases as PFF treatment time increases. (4) Glb1l2 expression decreases after D3 of PFF treatment, then increases again as PFF treatment time increases.

### PD progression GWAS SNPs that are correlated with TRIM2 expression changes are validated in additional genetic cohorts

To further prioritize, we next hypothesized that the SNPs identified by our PD Progression GWAS might regulate gene expression in human brain tissue. GTEx eQTL analysis in 13 brain regions revealed that at least three PD progression GWAS SNPs are associated with expression changes of *GLB1L2* and *TRIM2* (**Figure 3a, Supplementary Table 8**). Rs147142161 was correlated with *GLB1L2* expression level in the anterior cingulate cortex (P=0.04). Rs72729647 and rs62323742 were associated with *TRIM2* expression levels in multiple CNS regions (**Supplementary Figure 3**), including the cortex and spinal cord (cervical c-1) for rs72729647 (P=0.004-0.005) and the cortex, hypothalamus, spinal cord (cervical c-1), and substantia nigra for rs62323742 (P=0.001-0.03). No colocalization signal between GWAS and brain eQTL data was observed in this locus (**Supplementary Note**). While other PD progression GWAS SNPs may also regulate the expression levels of their corresponding genes, their eQTL status cannot be readily validated due to their rare appearance in population. Overall, brain eQTL data show that our PD progression GWAS SNPs correlate with changes in *TRIM2* expression levels in multiple PD-relevant brain tissues.

**Figure 3:**
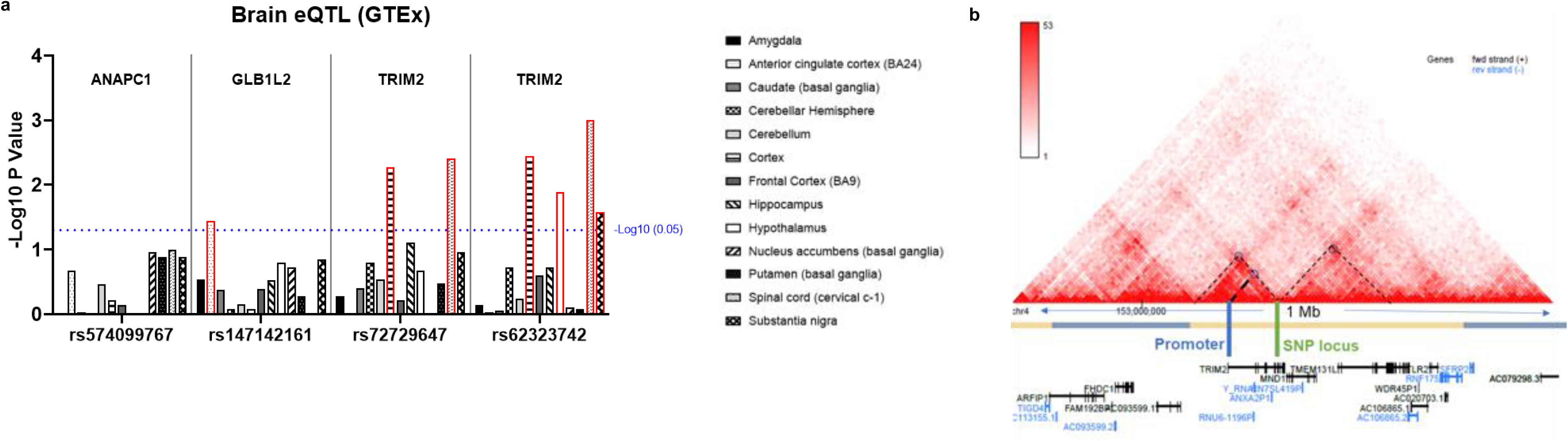
Our PD progression GWAS SNPs are associated with TRIM2 expression level and promoter activity. **a.** GTEx brain eQTL data of PD progression GWAS SNPs for variants with known gene annotations in the locus. Dotted blue line denotes statistical significance cutoff for P value <0.05. Tissues with statistically significant p values are marked in red. **b**. Chromosomal conformation data reveals topological association between the PD progression GWAS SNP locus and the TRIM2 promoter region in the brain (glioblastoma).

Brain chromosomal conformation data further revealed a strong physical interaction between the two GWAS SNPs and the *TRIM2* promoter region, suggesting that these SNPs regulate TRIM2 transcription (**Figure 3b, Supplementary Note**)^20^. Further, genetic variants that are linked to the two GWAS SNPs reside in open chromatin marks, suggesting potential regulatory roles for the associated *TRIM2* variants (**Supplementary Figure 4, Supplementary Note**).

We next investigated whether the *TRIM2* PD progression GWAS SNPs influenced measures of clinical progression in additional human PD cohorts. Importantly, the *TRIM2* PD progression GWAS SNPs showed the same directional effects in multiple PD progression phenotypes in two other datasets (**Supplementary Note**). In the Iwaki *et al.* GWAS meta-analysis dataset (12 longitudinal cohorts), the two SNPs are associated with three PD motor progression phenotypes (**Table 2)**^21^. Specifically, (1) UPDRS3_scaled (motor examination) was significantly associated with rs72729647 (P=0.009) and rs62323742 (P=0.006), (2) time to motor fluctuation was significantly associated with rs62323742 (P=0.01), and (3) UPDRS4_scaled (motor complication) was weakly associated with rs72729647 (P=0.06). Further, in the Genomics England PD progressor dataset, which was derived from the electronic medical records and WGS dataset of 100,000 UK individuals (unpublished, N=193), the same directional effects were observed for both SNPs (**Supplementary Table 8)**, when comparing the genotypes of the PD fast progressors (FP) and the not-fast progressors (NFP): rs72729647 minor allele present in 41% and 33% of FP and NFP respectively (P = 0.29), and rs62323742 minor allele present in 62% and 50% of FP and NFP respective (P = 0.11)^22^. Such repeated directional effects of the *TRIM2* SNPs on PD progression, despite the different PD progression phenotype definitions and genetic heterogeneity, provide strong validation for these SNPs’ association with PD progression, which may be replicated at the genome-wide level when larger longitudinal PD cohorts with deep phenotype information become available.

**TABLE 2.** TRIM2 PD progression SNPs in Iwaki *et al*. dataset.

| Phenotype analyzed in meta-analysis | # Individuals (# Datasets) analyzed | rs72729647 p-value | rs62323742 p-value |
| --- | --- | --- | --- |
| UPDRS3_scaled (motor examination) | 932 (4)* | **0.009** | **0.006** |
| UPDRS4_scaled (motor complication) | 254 (2) | 0.06 | 0.1884 |
| Time to motor fluctuation | 557 (2)* | 0.182 | **0.01** |

\*Potential overlap with PPMI individuals

### Multiple evidence suggests TRIM2 as a candidate for functional validation

Due to the combined genetic, transcriptomic, and regulatory evidence above, as well as the brain- specific expression of TRIM2 and its known cellular and clinical functions, we decided to investigate a potential role for TRIM2 function in PD^23^. We viewed these follow-up validation studies to be particularly important due to the relatively small sample size (N=319) of the PPMI cohort from which this PD progression GWAS candidate was identified. More specifically, we pursued TRIM2 functional validation due to the following 5 factors: (1) the top two SNPs in our PPMI PD progression GWAS were both intronic variants of *TRIM2*; (2) the eQTL, chromosomal conformation, and enhancer data suggest that the GWAS SNPs may regulate *TRIM2* expression in PD-relevant CNS tissues; (3) the effect of these SNPs on motor progression phenotypes were corroborated in two additional PD datasets while other GWAS SNPs in our dataset could not be validated due to their low population frequencies; (4) rare *TRIM2* loss-of-function mutations cause Charcot-Marie-Tooth disease type 2R (CMT2R), an axonal peripheral neuropathy with various motor impairments (OMIM:615490); and (5) the known role for TRIM2 in regulating axonal NF- L accumulation via ubiquitination permits a possible cogent molecular mechanism by which changes in TRIM2 expression might impact PD pathophysiology and progression^24,25^. The last point is noteworthy since NF-L has long been known to co-accumulate with α-synuclein in Lewy bodies^26,27^.

### TRIM2 regulates the accumulation of neurofilament-L in neurons

To investigate the impact of TRIM2 on cellular phenotypes of PD progression, two experimental systems were established in mouse primary neurons: Trim2 siRNA knockdown and lentivirus (LV) overexpression. We confirmed that the treatment of mouse Trim2 siRNAs led to >90% reduction in Trim2 mRNA and protein levels, while neurons transduced with LV expressing human TRIM2 exhibited ∼2-fold increase in TRIM2 protein (**Supplementary Figure 5a-d**). We also showed that TRIM2 knockdown and overexpression increased and reduced total levels of neurofilament-L (NF-L), respectively (**Supplementary Figure 5e-f**), consistent with the known role for TRIM2 in ubiquitination and proteasomal degradation of NF-L^24,25^.

Since NF-L is known to colocalize with LBs in PD, we hypothesized that NF-L would also colocalize with α-syn pathology in cellular models of PD^26–28^. To test this hypothesis, we first treated mouse primary neurons with PFFs to induce α-syn aggregation. PFF treatment (14d) induced robust formation of pS129+ α-syn aggregates, a surrogate marker for *de novo* α-syn aggregate formation, in the neurites and soma (**Supplementary Figure 6a-d**). NF-L immunostaining revealed that ∼20% of pS129+ α-syn aggregates were also positive for NF-L, confirming that NF-L accumulates with a subset of α-syn aggregates (**Figure 4a-b**). We also observed a strong correlation (R^2^=0.9191, P=0.0007) between the levels of NF-L and pS129+ α- syn accumulation in neurons treated with PFFs (**Supplementary Figure 6e**). Treatment with TRIM2 siRNAs further revealed that TRIM2 reduction increases both total NF-L accumulation (**Figure 4c**) and the colocalization of NF-L with pS129+ α-syn aggregates in the neuronal soma (**Figure 4d**), while maintaining the overall percentage of pS129+ α-syn aggregates co-positive for NF-L (**Figure 4b**).

**Figure 4:**
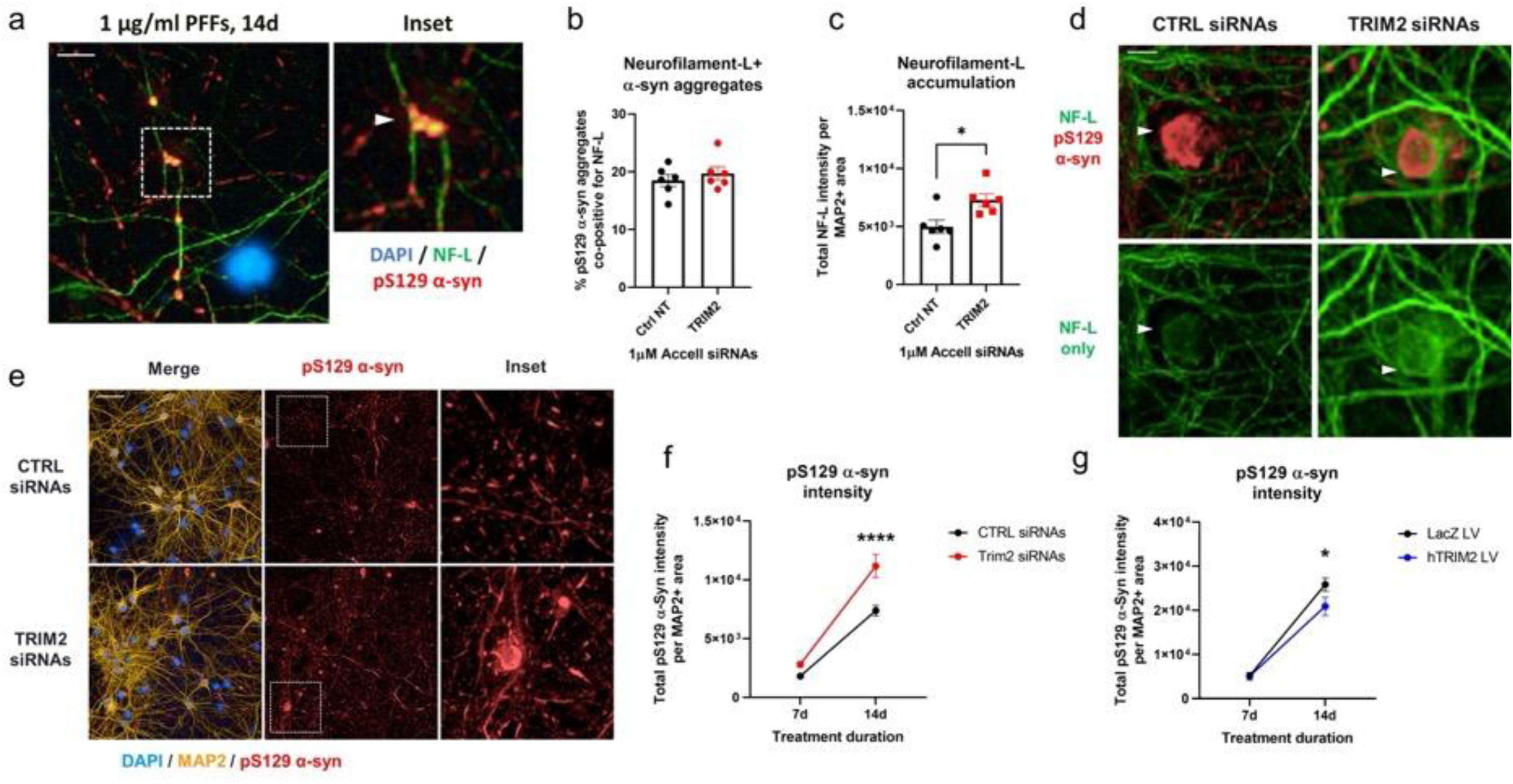
TRIM2 regulates the accumulation of neurofilament-L and pathological α-synuclein. CD1 mouse primary neurons were cultured for 21 days in vitro (DIV) with or without Accell siRNA knockdown of murine TRIM2 and treated with 1 µg/ml human α-syn preformed fibrils (PFFs) for the final 7 or 14 days. **a.** Representative images of DIV 21 neurons treated with a-syn PFFs for 14d and stained for DAPI (blue), neurofilament-L (NF-L, green), and pS129 α-syn (red). Arrows denote inclusions that are co-positive for NF-L and pS129 α-syn. Scale bar = 10µm. **b-c.** Quantification of the percentage of pS129 a-syn aggregates that are co-positive for NF-L (b) and total NF-L intensity per MAP2+ area (c) in a. *p=0.0141 by unpaired Student’s t-test; n=6wells/group. **d.** Representative images of neuronal cell body pS129 α-syn aggregates (red, arrows) and NF-L (green, bottom panel) in neurons treated with 1 µg/ml PFFs for 14d. Note the accumulation of NF-L in neurons treated with TRIM2 siRNAs. Scale bar = 5µm. **e**. Representative images of DAPI (blue), MAP2 (orange), and pS129 α-syn (red) immunostaining in DIV 21 CD1 neurons treated with TRIM2 or non-targeting control (CTRL) siRNAs and 1 µg/ml PFFs for 14d. Scale bar = 20µm. **f-g.** Quantification of total pS129 α-syn pathology per MAP2+ area in (e). after 7d or 14d treatment with PFFs for CD1 neurons treated with TRIM2 siRNAs (f) or human TRIM2 lentivirus (hTRIM2 LV, g). ****p<0.0001 *p=0.0313 by 2-Way ANOVA with Sidak’s multiple comparisons test; n=9 wells/group. Figures a-g include data from a single representative experiment repeated at least 3 times in independent cultures of primary mouse neurons. Error bars represent mean ± SEM.

### TRIM2 regulates accumulation of pathological α-synuclein

We next tested whether TRIM2 protein levels had a direct impact on the accumulation of pS129+ α-syn pathology. For this, we used primary neurons cultured from two mouse lines: CD1, expressing wildtype murine α-syn at endogenous levels, and M83, overexpressing human A53T mutant α-syn. M83 neurons exhibit accelerated α-syn aggregation compared to CD1 in response to PFF treatment, and reliably develop pS129+ α-syn aggregates in the cell body of neurons that are reminiscent of Lewy body pathology in PD within 7-14 days of PFF treatment (referred to as ‘Lewy-like’ pS129+ α-syn in this manuscript) (**Supplementary Figures 7-8**).

TRIM2 knockdown significantly increased the percentage of M83 neurons containing Lewy-like aggregates (**Supplementary Figure 9a**). This was also reflected in the quantification of total cell body pS129+ α-syn aggregate area and intensity (**Supplementary Figure 9b-c**), without any impact on neuronal viability in control or PFF-treated cells (**Supplementary Figure 9d-e**). TRIM2 knockdown in CD1 neurons also resulted in increased accumulation of pS129+ α-syn pathology between 7-14 days (**Figure 4e-f**).

Overexpression of TRIM2 in CD1 neurons resulted in reduced pS129+ α-syn pathology (**Figure 4g**). Analysis of fibril uptake with pHrodo Red-conjugated α-syn PFFs confirmed that the observed effect of TRIM2 protein level on α-syn aggregation could not be explained by differences in internalization of α-syn PFFs by TRIM2 knockdown or TRIM2 overexpressing neurons (**Supplementary Figure 9f-g**); TRIM2 knockdown and overexpression also did not impact levels of total soluble α-syn in neurons (**Supplementary Figure 5a**). These results demonstrate that TRIM2 level regulates neuronal α-syn aggregation.

Apart from α-syn aggregation, TRIM2 knockdown impacted additional PD-associated cellular phenotypes. TRIM2 knockdown significantly reduced activity of the lysosomal enzyme β-glucocerebrosidase (GBA, or GCase, **Supplementary Figure 10a-c**), which is also observed in normal aging and PD progression^29^. GBA mutations are the most prevalent risk factor for PD and it has been suggested that the carriers exhibit a faster progression of motor and cognitive symptoms in PD^5,6,30^. Additionally, TRIM2 reduction induced an increase in mitochondria number and a decrease in mitochondria size as measured by MitoTracker, suggesting an overall fragmentation of the mitochondrial network (**Supplementary Figure 11a-d**). Mitochondrial dysfunction is implicated in PD through both environmental and genetic factors, including familial PD genes such as *PINK1* and *PRKN*^31^. Our results suggest that loss of TRIM2 function may contribute to mitochondrial dysfunction. However, whether TRIM2 regulates mitochondrial respiration or turnover in response to oxidative stress or damage remains unknown.

### *TRIM2* GWAS SNPs correlate with peripheral NF-L levels in PD patients

Finally, we examined whether the *TRIM2* GWAS SNPs impacted peripheral biomolecules that are associated with PD progression via comprehensive peripheral multi-omic (proteome, transcriptome, methylome) analyses (**Supplementary Note**). Indeed, *TRIM2* GWAS SNP carriers exhibited increased NF-L levels in plasma and CSF (**Supplementary Figure 12).** Specifically, rs72729647 was correlated with higher CSF NF-L level (P = 0.007, N=64, **Figure 5a**), and rs62323742 was correlated with higher plasma NF-L level (P = 0.06, N=71, **Figure 5b**). This peripheral biomolecule data supports a potential role for TRIM2 dysfunction in PD. However, whether the higher levels of NF-L in plasma and CSF in *TRIM2* variant carriers are a direct result of loss of TRIM2 function, or an indirect effect of the neuronal damage associated with accelerated α-syn pathology remains unknown.

**Figure 5:**
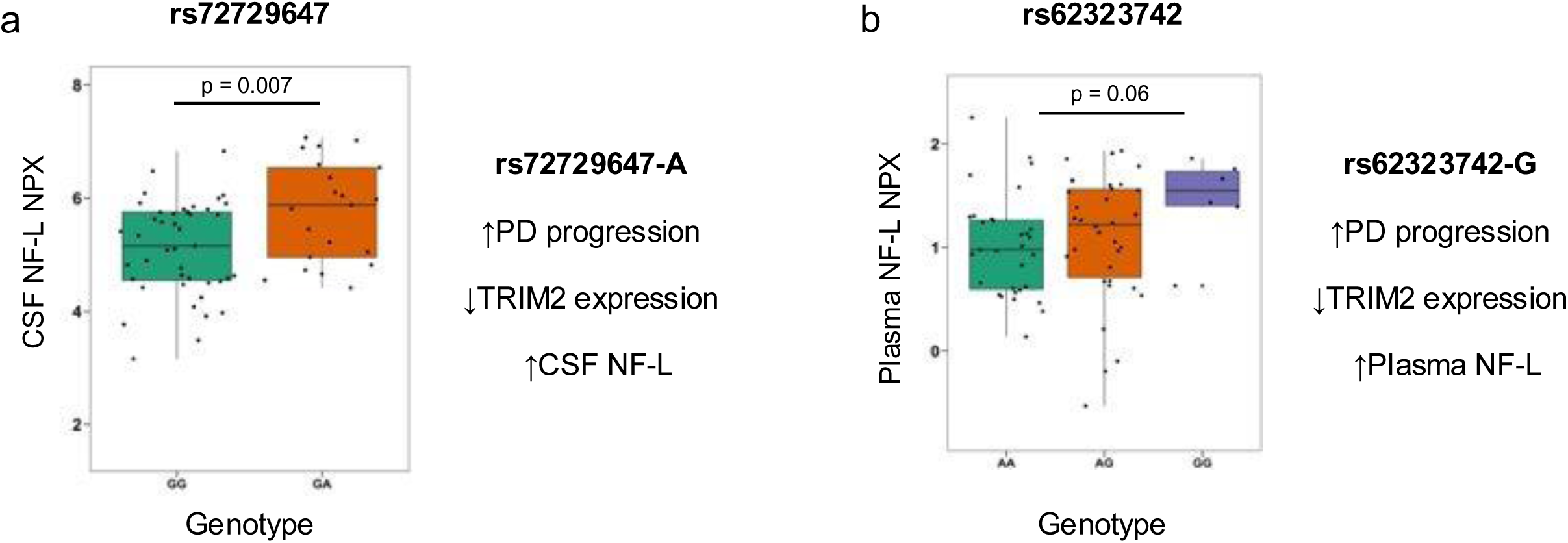
Progression SNPs in TRIM2 are associated with increased neurofilament-L (NF-L) in patient biofluids. **a-b.** Peripheral NF-L protein levels in PD patients (PPMI and PDBP) stratified by TRIM2 GWAS variants, rs72729647 (a) and rs62323742 (b). Homozygous for reference allele (green), heterozygous (orange) and homozygous for alternative allele (purple). **p=0.007 (GG vs. GA) *p=0.06 (GG vs. AA) by Welch’s t-test.

## DISCUSSION

Here, we integrate the results of two new PD progression GWAS with omics data from PD-relevant cellular models to identify candidate genes underlying PD progression. We identified 8 protein-coding genes as candidates for modifiers of PD progression at the genome-wide significant level using longitudinal clinical ratings of 1,433 PD patients. While none of these have been genetically associated with PD before, all of them are expressed in the brain, with some particularly relevant to PD. *TRIM2* and *ANAPC1* encode E3 ubiquitin ligases, the same protein family as parkin (PRKN) that causes familial PD when mutated^23,32^. This protein family plays a critical role in the ubiquitination process that degrades proteins and damaged organelles via the ubiquitin proteasome and autolysosomal systems, respectively. Impairments in the ubiquitination system are implicated in PD, leading to reduced clearance of intracellular protein aggregates and damaged mitochondria^33^. *ZBTB20* and *MED13L* encode transcription factors that regulate neurogenesis^34^. ZBP1 regulates necroptosis induced by TNF, which is implicated in PD pathophysiology via neuroinflammation^35^. PPP2R3A is active in mitochondria, whose dysfunction is implicated in PD etiology^29,36^. Furthermore, *Ppp2r3a* -/- mice exhibit decreased grip strength and rare variants in *PPP2R3A* have been associated with PD^37,38^. GLB1L2 is specifically enriched in rod photoreceptor cells in retina, where the disturbance is often observed in early PD^39,40^. Importantly, four genes (*TRIM2, MED13L, PPP2R3a, GLB1L2*) were also identified as PFF induced-DEGs in a cellular model of PD, suggesting that these genes are involved in α-syn aggregation or the cellular response to neuropathology. Further, our comparisons of tentative GWAS signals and PFF-induced DEGs produced a comprehensive list of 137 genetic candidates for PD progression, potentially suitable for high-throughput functional screening (**Supplementary Note**).

The GWAS and omics analyses presented in our study suggest several new candidate genes and pathways associated with the clinical progression of PD. However, our approach towards defining PD progression has potential limitations. First, the results may be partially confounded by potential effects of age on PD progression. Yet, given that PD is an aging-associated disease, removing aging effect from disease progression would likely lead to overly conservative results, missing many true associations^37,38^. Second, PD progression may be influenced by disease duration, which is difficult to accurately compute due to lack of diagnostic biomarkers for PD^41^. This may be of particular concern for our AMP-PD data, where multiple cohorts with different protocols are combined, as it is challenging to ensure that the patients’ clinical and molecular measurements at the baseline are representing the same disease stage. In PPMI, where the protocol strictly enrolls early PD patients only, this may be less of concern. Third, our GWAS model does not incorporate possible changes in PD progression rate over the course of PD. This may have contributed to the non-replicating PPMI GWAS result in AMP-PD data, where PD progression rate was calculated for substantially longer duration than in PPMI dataset, in addition to the genetic and phenotypic heterogeneity. Lastly, while our quantitative GWAS design provides superior statistical power to conventional case-control design, the sample sizes are still relatively small. Continuing efforts to expand longitudinal cohort with well-controlled clinical measurements will further define the genetic landscape of PD progression.

Due to the potential limitations above, we viewed functional validation of the candidate PD progression GWAS genes to be particularly important. Follow-up analysis of the 8 PD progression GWAS candidate genes that reached genome-wide significance revealed that the GWAS SNPs in the *TRIM2* locus were likely to impact TRIM2 expression level in several brain tissues relevant to PD. We therefore hypothesized that TRIM2 expression level would also impact α-syn aggregation in cellular models of PD. Our results indicate that TRIM2 knockdown in mouse primary neurons accelerates α-syn aggregation in response to PFF treatment. Conversely, overexpression of TRIM2 reduces PFF-induced α-syn pathology. Our results also confirmed that TRIM2 expression level regulates NF-L accumulation in neurons, which was associated with the extent of α-syn aggregate formation. Analysis of human CSF and plasma samples from PD patient cohorts confirmed that the PD progression GWAS SNPs in the *TRIM2* locus are associated with differential levels of NF-L. While peripheral NF-L levels have been validated as a biomarker of neuronal damage in other neurodegenerative diseases, the role of NF-L as a biomarker in PD is unknown. Therefore, the increased NF-L levels observed in *TRIM2* SNP carriers (**Figure 5**) may be the result of more pronounced neurodegeneration in these patients, which would be consistent with the increased rate of PD progression, or a TRIM2-mediated effect that is unrelated to the effect of the SNPs on disease progression. Further research is needed to elucidate the mechanism by which the GWAS SNPs in the *TRIM2* locus impact both TRIM2 function and disease progression.

Based on the currently available data from human patients and mouse neuron experiments presented here, we propose the following molecular model for TRIM2 involvement in PD progression (**Figure 6**): NF-L and α-syn both accumulate in PD, with a significant portion of α-synuclein and NF-L aggregates co-localizing in neurites and the cell body. TRIM2 ubiquitinates NF-L, thereby targeting it for proteasomal degradation and promoting normal protein homeostasis. Loss of TRIM2 function results in aberrant accumulation of NF-L, which may serve to further promote α-syn aggregation, inhibit α-syn clearance by other mechanisms, or both. Conversely, activation of TRIM2 activity or upregulation of TRIM2 expression may slow the progression of PD by both reducing α-syn pathology and promoting normal protein homeostasis in neurons. In addition to α-syn pathology, TRIM2 may impact PD progression via the regulation of lysosomal and mitochondrial activities. While future studies are warranted to understand the exact role of TRIM2 in mediating the clinical progression of PD, activation of the TRIM2 pathway may represent a novel therapeutic approach for disease modification in PD.

**Figure 6:**
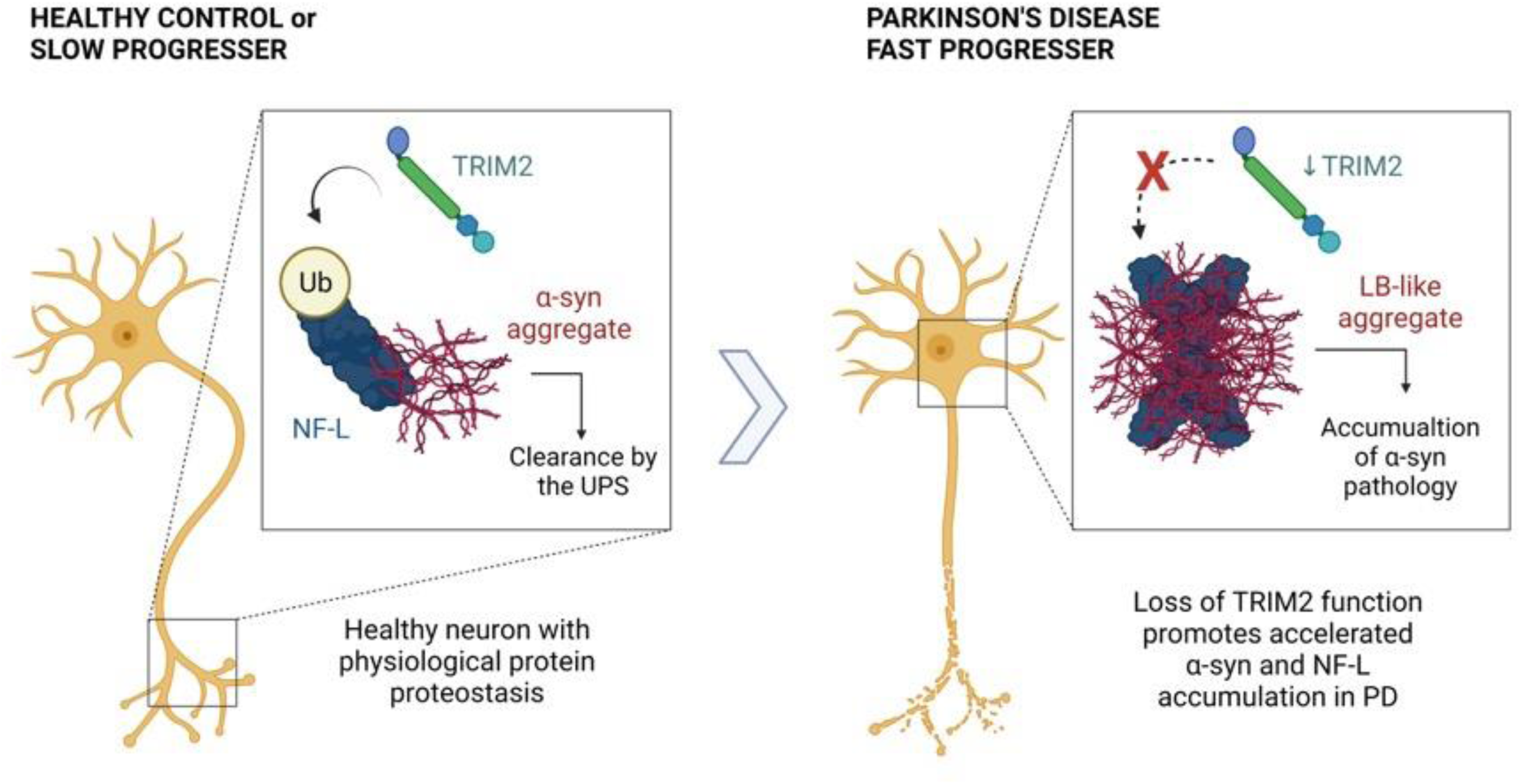
Model for TRIM2 involvement in PD progression. Neurofilament-L (NF-L) and α-syn both accumulate in PD, with a significant portion of α-syn and NF-L aggregates co-localizing both in neurites and in the cell body. TRIM2 ubiquitinates NF-L protein, thereby targeting it for proteasomal degradation and promoting normal protein homeostasis (left panel). Loss of TRIM2 function results in aberrant accumulation of NF-L which may serve to further promote α-syn aggregation, inhibit α-syn clearance by other mechanisms, or both (right panel). Therefore, activation of TRIM2 activity or upregulation of TRIM2 protein level may slow the progression of PD by both reducing α-syn pathology and promoting normal cellular homeostasis in neurons.

Our study integrates patient data from longitudinal clinical PD cohorts with cellular models of PD to identify potential therapeutic targets for modifying PD progression. Future studies on these candidate genes may identify biological mechanisms that are tractable for therapeutic intervention in PD.

## Online Methods

### AMP-PD WGS data

Whole genome sequence (WGS, hg38), demographic data and clinical scores of 3,359 PD patients were obtained via tier 2 access to the AMP-PD v2 December 2020 release. The sequencing data was generated by the Illumina HiSeq X Ten sequencer with samples coming from whole blood. Variants were called using GATK Best Practice. The final analyzed data comprised of 1,114 PD patients from 3 cohorts (PPMI, PDBP and STEADY-PD III (Safety, Tolerability, and Efficacy Assessment for Isradipine for PD)) and 8,762,817 autosomal genetic variants were retained after filtering for the following criteria: (1) include PD patients with MDS-UPDRS total score measured for at least 2 time points, (2) exclude self-reported non-white individuals, (3) exclude samples and variants with genotype missingness rate of >0.1, (4) include variants with allele count (AC) > 10 and QUAL = PASS only.

### PPMI WGS data

We obtained 960 individuals’ WGS from the PPMI. The dataset was generated by Illumina HiSeq XTen sequencing and variant calling using GATK Best Practice, aligned in hg38. The final analyzed set included 319 PD patients and 46,853,470 autosomal genetic variants, after applying the following criteria: (1) include the PD patients with MDS-UPDRS total score measured for at least 2 time points, (2) exclude self-reported non-white individuals and phenotype outliers, (3) exclude samples and variants with genotype missingness rate of >0.1, and (4) include variants with QUAL=PASS only. For downstream analyses and visualization, variants with AC>2 were included.

For clinical and demographic data, the curated data cuts for the original cohort (423 newly diagnosed PD patients and 196 age-matched healthy controls) including the baseline and Year 1-3 were downloaded from the PPMI LONI data portal on 5/16/2019.

### GWAS

For each patient, MDS-UPDRS total score (sum of part I, II and III), was obtained or calculated for each visit^42^. The annual rate of change in each patient’s score was then defined as the slope calculated from linear regression of the MDS-UPDRS total scores over time. The obtained ‘PD progression rate’ was treated as quantitative trait (y) in the linear model for association analysis.

To avoid spurious associations, potential confounders were assessed. Sex was used as covariate in both GWAS analyses. Other potential factors, including genetic relatedness, population structure, and clinical sites were assessed for each dataset and incorporated as covariates when necessary (**Supplementary Note**).

The GWAS loci were defined as taking the furthest genetic variants linked to its lead variant (r^2^>0.5, ±1Mb) and merging overlapping/nearby intervals (<250kb).

Genetic associations with p < 5E-8 were considered genome-wide significant, and those with p < 1E-5 were categorized as those with suggestive associations. Association analysis and genetic data assessment were performed using *PLINK 1.9* and *2.0*. R packages *ggplot2*, *dplyr*, *qqman*, and *gap* were used for data visualization.

### Aggregate variant analysis

PPMI WGS variants were annotated with predicted consequences using VEP^43^. SKAT-O test was performed in 4 categories: (1) loss-of-function (LoF) only, (2) LoF with rare variant weight, (3) LoF + missense, (4) LoF + missense with rare variant weight. For LoF category, variants predicted as frameshift, start lost, stop gained were included. For LoF + missense category, variants predicted with inframe deletion, inframe insertion, missense predictions were included, in addition to LoF variants. The same covariates as PPMI GWAS were used, and rare variant weight was applied using default beta (1,25). Following variants and tests were excluded: (1) multi-allelic sites, (2) variants with AC<2, (2) test with only 1 variant site, (3) test with <5 cumulative AC. Bioconductor package GENESIS was used to perform SKAT-O.

### Pathway analyses

Pathway and Gene Ontology (GO) enrichment analyses were performed using gprofiler2, unless otherwise noted^44^. For analyses on mouse genes, mouse genes were first converted to human orthologs using *getLDS* function implemented in *biomaRt*^45^. Enrichment analyses with default background list, was performed for each gene list against KEGG, Reactome, and GO databases, and only results with adjusted p-value < 0.05 were considered significant.

### PFF-induced transcriptome

We downloaded the publicly available bulk mouse primary neuron RNA-seq data (GSE142416)^8^. The RNA-seq reads were aligned to the mm10 mouse reference genome with STAR v2.5.3a using the parameters --alignSJDBoverhangMin 1, --outSAMstrandField intronMotif, -- outFilterMismatchNoverLmax 0.1, and --alignIntronMax 1000000, and using samtools v1.6.0^46,47^. We used featureCounts from subread v1.6.0 for feature counting with the parameters -p, -s 2, -C, and --minOverlap 1^48^. Picard v2.18.7 was used for quality control (QC): We looked at proportion of reads mapping to coding bases vs. other bases, normalized coverage along the gene body of the 1000 most highly expressed transcripts, distribution of expression signals, and PCA. As a result, no samples were excluded. We kept any genes with counts-per-million (cpm) greater than 1 in at least 3 samples. We used trimmed mean of M-values (TMM) to normalize the raw counts, and *limma-voom* (*limma* v. 3.44.3) for differential expression analysis of pairwise comparisons for the timepoints^49^. Datasets from neurons treated with PBS at DIV5 were used as controls.

For pathway analyses, we first converted genes from mouse to human using the Ensembl mart in biomaRt^45^. We used *clusterProfiler* v3.16.1, which applies hypergeometric tests^50^. We used the list of all genes tested for differential expression as a background list and all genes with FDR < 0.05 and absolute logFC >= 1 as input. A pathway/gene set was tested if at least five genes were present in the background list. We considered gene sets and pathways with adjusted p-values < 0.05 as significant. We performed enrichment analyses on results from all pairwise comparisons except for D1 vs. PBS and D14 vs. D7 which only had 31 and 29 input genes, respectively.

### TRIM2 variant eQTL

For targeted eQTL analysis, pre-computed brain eQTL data from 13 brain regions were obtained from the GTEx Portal. All four genes within 500Kb of our TRIM2 candidate variants were evaluated for their nominal eQTL p-values and the normalized transcript expression levels by genotypes.

### Colocalization Analysis

Colocalization analyses were performed to identify variants that affect PD progression in the brain. For the PD progression trait, summary statistics generated from our AMP-PD and PPMI GWAS were used. For transcription as a trait, GTEx v8 brain expression quantitative trait loci (eQTLs) from 13 different brain regions, and PD eQTLs mapped using PPMI genotype and baseline gene expression data were used^23,51^. *coloc* R package was used to calculate the posterior probabilities that a variant affects both traits^52^.

### GeL and Iwaki datasets

The GeL PD progression dataset was derived from the Genomics England (GeL) 100,000 genome project. GeL includes 107,513 WGS of patients with rare disease or cancer in England, linked to secondary care data from Hospital Episodes Statistics (HES). 81 PD fast progressor (FP) and 112 not-fast progressor (NFP) were defined based on the combinations of the longitudinal records of ICD-10 codes and medication prescriptions. The manuscript describing our algorithm is in submission (Noh *et al*.). For the calculation of *TRIM2* candidate variant carrier frequency in FP and NFP groups, homozygote and heterozygote carriers were considered equally.

We obtained the GWAS summary statistics of the Iwaki dataset^4^. This dataset includes GWAS meta-analysis of 25 phenotypes in 4,093 PD patients from 12 longitudinal cohorts with median follow-up duration of 3.81 years. We interrogated the p-values of our TRIM2 variants for all 25 phenotypes reported in the study.

### Primary neuronal cell culture

Primary neuronal cultures were generated by dissection of hippocampal and cerebral cortex tissues from embryonic day 16 (E16) mice according to AbbVie IACUC approved protocols. Briefly, tissues were dissected in ice-cold HBSS (Invitrogen, Cat# 14175095), separated from the meninges, and washed 3x in HBSS on ice. Tissues were then dissociated by incubation with neuronal isolation enzyme (ThermoFisher Scientific, Cat# 88280) at 30°C for 30min; the tubes were gently inverted every 5min to enable efficient enzyme digestion. After 30min, the tissues were gently triturated by manual pipetting 5-10x, resuspended in 8ml of plating media (DMEM [Invitrogen, Cat# 11960044] supplemented with 10% fetal bovine serum, 1x penicillin-streptomycin), and filtered using a 70µm cell strainer (Falcon, Cat# 087712). Cells were then counted and plated in 150ul plating media per well in poly-D-lysine pre-coated 12- or 96-well plates (Corning, Cat# 353219). 4h later, plating media was aspirated and replaced with neuron maintenance media (Neurobasal Plus [Invitrogen, Cat# A3582901] supplemented with B-27 Plus [Invitrogen, Cat# A3582801], 1x penicillin-streptomycin, and 1x GlutaMAX [Invitrogen, Cat# 35050061]). 1/3 volume fresh neuronal maintenance media exchange was performed twice weekly until day of analysis.

### α-syn PFF seeding assays

Wildtype full-length human α-syn PFFs were generated as described previously and obtained from the Luk lab at the University of Pennsylvania^14^. PFFs were diluted to 0.1mg/ml in Dulbecco’s PBS (DPBS) and sonicated for 1 min at 30% amplitude with 1s on, 1s off pulses using a QSonica water bath sonicator. Sonicated PFFs were added directly to the extracellular media with fresh neuronal maintenance media on either day in vitro (DIV) 7 or 14. Cells were treated for 14d or 7d, respectively, and fixed for analysis on DIV 21. For phosphor-serine 129 (pS129) α-syn immunocytochemical analysis, cells were fixed in ice-cold methanol at –20C for 20min followed by 3x washes in DPBS. Cells were stored at 4C in DPBS until the day of staining.

### Immunocytochemistry (ICC)

Cells were rinsed 3x in DPBS followed by permeabilization in DPBS buffer containing 0.1% Triton X-100 for 20 min. Cells were then incubated in block buffer containing 5% normal goat serum and 0.05% Triton X-100 diluted in DPBS for 1h. After blocking, cells were incubated in primary antibodies diluted in primary antibody incubation buffer (DPBS containing 2% bovine serum albumin and 0.05% Triton X-100) overnight at 4°C. Primary antibodies used for ICC include: pS129 α-syn [D1R1R] rabbit mAb (Cell Signaling Technology, Cat# 23706, 1:3000), MAP2 chicken antibody (abcam, Cat# ab92434, 1:5000), anti-NeuN mouse mAb, [clone A60] (Millipore, Cat# MAB377, 1:2000), and Neurofilament-L [DA2] mouse mAb (Cell Signaling Technology, Cat# 2835, 1:2000). The next day, the cells were washed 3x with PBS-T wash buffer (DPBS/0.05% Triton X-100), 10 min each, followed by incubation in secondary antibodies conjugated with Alexa 488, Alexa 568, and Alexa 647 fluorophores for 2h. Secondary antibodies were diluted in block buffer supplemented with 1ug/ml DAPI. After 2h incubation with secondary antibodies, cells were washed 3x with PBS-T, 10 min each, and stored in DPBS at 4°C until imaging using the Perkin Elmer Opera Phenix high-content confocal microscope system. Unless stated otherwise, all ICC steps were performed at room temperature with gentle agitation.

### Plasmids, RNAi, and lentiviruses

Lentiviral plasmids described in this study were purchased from The Broad Institute Functional Genomics Center (FGC). LacZ control and human full-length TRIM2 constructs were driven from the ubiquitous EF1a promoter in an pLX_TRC317 vector. Lentivirus (LV) production and titer determination was performed by Alstem. Unless noted otherwise, LVs were added to primary neurons with fresh neurobasal maintenance media on DIV 4 at a multiplicity of infection of 2 genome copies per cell.

Accell transfection-free siRNAs were purchased from Dharmacon (cat# D-001910-10-50, E-047102-01-0010) and added to primary neurons with fresh neurobasal media at a concentration of 1 uM on DIV 4.

### Protein extraction and Western blotting

Lysates for biochemistry were prepared from neurons in 12-well plates (500,000 cells per well). Briefly, neurons were lysed in RIPA lysis buffer (ThermoFisher Scientific, Cat# 89900) supplemented with Halt protease and phosphatase inhibitor cocktail (Pierce, Cat# 78443), centrifuged at 10,000rpm for 15min at 4C, and the supernatants were removed to determine protein concentration by BCA protein assay kit (Pierce, Cat# 23225). Samples were prepared for Western blot using LDS sample buffer and separated by gel electrophoresis using Bolt 4-12% Bis-Tris SDS-PAGE gels. Proteins were transferred to nitrocellulose membranes using the iBolt semi-dry transfer system and membranes were blocked in 5% non-fat dry milk (NFDM)/TBS-T (0.05% Tween-20) for 1h. Primary antibodies were diluted in 2% BSA/TBS-T solution and incubated with membranes overnight at 4C. Antibodies used for Western blot include: rabbit anti-TRIM2 pAb (Proteintech, Cat# 20356-1-AP; 1:1,000 dilution), mouse anti-Neurofilament-L [DA2] mAb (Cell Signaling Technologies, Cat# 2835S; 1:1,000 dilution), rabbit anti-α-synuclein [MJFR1] mAb (Abcam, Cat# ab138501; 1:,5000 dilution), and mouse anti-ACTB (Abcam, Cat# ab8226, 1:5,000). After primary antibody incubation, membranes were washed 3x with TBS-T, 10 min each followed by incubation in HRP-conjugated secondary antibodies diluted 1:10,000 in 1% NFDM/TBS-T for 1h at room temperature. After secondary antibody incubation, membranes were washed 3x in TBS-T for 10 min each. Immunoblots were developed and imaged using ECL substrate solution (ThermoFisher Scientific, Cat# 34096) and a BioRad chemilluminescence imager.

### Live-cell imaging

For live-cell lysosomal GCase activity assays, neurons were incubated with 5 nM MDW941 activity-based probe, 50 nM LysoTracker Deep Red (Invitrogen, Cat# L12492), and NucBlue Hoechst nuclei stain (Invitrogen, Cat# R37605) for 2h at 37C. MDW941 probe was synthesized by WuXi according to the chemistry scheme described by Witte *et al*^53^. MDW941 positive fluorescence was then imaged at 37C in live cells. 500uM CBE pre-treatment for 1h was used as a negative control to define background fluorescence; CBE is an irreversible inhibitor of GCase enzymatic activity. For mitochondrial morphology assays, neurons were incubated with 50 nM MitoTracker Green (Invitrogen, Cat# M7514) and NucBlue for 1h followed by live imaging.

### Immunofluorescence Microscopy and Image Analysis

All fluorescence images were captured at 20x or 40x magnification using the Perkin Elmer Opera Phenix high-content confocal imaging system and analyzed using Harmony image analysis software. For quantification of Lewy-like pS129 α-syn aggregates in the neuronal cell body, Lewy-like aggregates were defined as pS129 α-syn immunostaining within NeuN+ cytoplasm with mean fluorescence intensity > 10,000 and sum area > 10um2. For mitochondrial morphology studies, mitochondria objects were defined as fragmented if the area < 1µm2, length < 1µm, and width:length ratio > 0.6. Conversely, tubular or elongated mitochondria objects were defined as area > 1µm2, length > 1µm, and width:length ratio < 0.6. All statistical analysis for cellular imaging experiments was performed using Graphpad Prism software.

### PPMI and PDBP targeted proteomics

Matched CSF and plasma proteomics data was obtained from 117 PD patients from the PPMI and Parkinson’s Disease Biomarkers Program (PDBP) cohorts via the tier 2 access to AMP-PD v2.5 May 2021 release. A total of up to 332 samples for at least 8 study visits over a 5-year period were included in the analyses. The targeted proteomics data was generated using the Proximity Extension Assay based Olink Explore 1536 panel, preprocessed and quality checked as previously described (https://amp-pd.org/data/targeted-proteomics-data). Normalized Protein Expression (NPX) values on four panels (cardiometabolic, inflammation, neurology, and oncology), along with clinical and demographics data including MDS-UPDRS scores were downloaded from the AMP-PD Google cloud storage repository.

For both CSF and plasma, proteins that failed QC were converted to missing values and those with values below limit of detection (LOD) were replaced with the LOD value. Proteins missing in >50% of samples and samples with >10% missing proteins were excluded from the analyses, resulting in the following sample sizes: 332 (CSF: cardiometabolic), 329 (CSF: inflammation), 332 (CSF: neurology), 327 (CSF: oncology), 354 (Plasma: cardiometabolic), 317 (Plasma: inflammation), 332 (Plasma: neurology) and 337 (Plasma: oncology), across 1,463 proteins in each biofluid.

To calculate the total MDS-UPDRS score, the off-score for part III was used whenever available. Using *limma*, multiple linear models were individually fitted on each protein in each panel with the total MDS-UPDRS score as the fixed effect and patient ID as the random effect to account for longitudinal repeated measures, with sex, race, and study as covariates. Differential protein abundance associated with a unit change in MDS-UPDRS score was calculated using the empirical Bayes statistic in *limma*, with the Benjamini-Hochberg corrected p-value of 0.05 as the threshold. Pathway overrepresentation analyses for differentially abundant proteins were carried out across the panels using GO, KEGG and Reactome databases within *gprofiler2* R package.

## Data Availability

Data used in the preparation of this article were obtained on 2021-09-09 from the Parkinson's Progression Markers Initiative (PPMI) database (https://www.ppmi-info.org/access-data-specimens/download-data), RRID:SCR_006431. For up-to-date information on the study, visit http://www.ppmi-info.org. Data used in the preparation of this article were obtained from the AMP-PD Knowledge Platform. For up-to-date information on the study, visit https://www.amp-pd.org. This research was made possible through access to data in the National Genomic Research Library, which is managed by Genomics England Limited (a wholly owned company of the Department of Health and Social Care). Any additional data produced in the present study are available upon reasonable request to the authors.

https://www.amp-pd.org

## ACKNOWLEDGMENTS

Data used in the preparation of this article were obtained on 2021-09-09 from the Parkinson’s Progression Markers Initiative (PPMI) database (https://www.ppmi-info.org/access-data-specimens/download-data), RRID:SCR_006431. For up-to-date information on the study, visit http://www.ppmi-info.org. PPMI – a public-private partnership – is funded by the Michael J. Fox Foundation for Parkinson’s Research and funding partners, including 4D Pharma, Abbvie, AcureX, Allergan, Amathus Therapeutics, Aligning Science Across Parkinson’s, AskBio, Avid Radiopharmaceuticals, BIAL, BioArctic, Biogen, Biohaven, BioLegend, BlueRock Therapeutics, Bristol-Myers Squibb, Calico Labs, Capsida Biotherapeutics, Celgene, Cerevel Therapeutics, Coave Therapeutics, DaCapo Brainscience, Denali, Edmond J. Safra Foundation, Eli Lilly, Gain Therapeutics, GE HealthCare, Genentech, GSK, Golub Capital, Handl Therapeutics, Insitro, Jazz Pharmaceuticals, Johnson & Johnson Innovative Medicine, Lundbeck, Merck, Meso Scale Discovery, Mission Therapeutics, Neurocrine Biosciences, Neuron23, Neuropore, Pfizer, Piramal, Prevail Therapeutics, Roche, Sanofi, Servier, Sun Pharma Advanced Research Company, Takeda, Teva, UCB, Vanqua Bio, Verily, Voyager Therapeutics, the Weston Family Foundation and Yumanity Therapeutics.

Data used in the preparation of this article were obtained from the AMP-PD Knowledge Platform. No funding to disclose. For up-to-date information on the study, visit https://www.amp-pd.org. The AMP® PD program is a public-private partnership managed by the Foundation for the National Institutes of Health and funded by the National Institute of Neurological Disorders and Stroke (NINDS) in partnership with the Aligning Science Across Parkinson’s (ASAP) initiative; Celgene Corporation, a subsidiary of Bristol-Myers Squibb Company; GlaxoSmithKline plc (GSK); The Michael J. Fox Foundation for Parkinson’s Research ; Pfizer Inc.; AbbVie Inc.; Sanofi US Services Inc.; and Verily Life Sciences. ACCELERATING MEDICINES PARTNERSHIP and AMP are registered service marks of the U.S. Department of Health and Human Services.

This research was made possible through access to data in the National Genomic Research Library, which is managed by Genomics England Limited (a wholly owned company of the Department of Health and Social Care). The National Genomic Research Library holds data provided by patients and collected by the NHS as part of their care and data collected as part of their participation in research. The National Genomic Research Library is funded by the National Institute for Health Research and NHS England. The Wellcome Trust, Cancer Research UK and the Medical Research Council have also funded research infrastructure.

We are grateful for the support of AbbVie GRC Bioinformatics Engineering, Computational Genomics, Human Genetics, and AbbVie Global Epidemiology teams, specifically: J. Tessmann, M. Reppell, and former AbbVie employees N. Chung and D. Oleske.

## AUTHOR CONTRIBUTIONS

H.J.N. conceived the project. H.J.N. designed the study with inputs from C.Z. and J.W.D.. A.S.K., L.G., J.W.D. and H.J.N. acquired datasets. D.A. and S.K. designed and conducted experiments with inputs from L.S., J.S. (Stender) and J.S. (Stoehr). A.S.K., D.A., S.T., L.G., L.N., R.L., S.G., S.L., Y.L., S.K., P.V. and H.J.N. analyzed data and interpreted results. A.S.K., D.A., S.T., L.G., L.N., S.G., and H.J.N. wrote the draft manuscript. All authors reviewed the manuscript and provided feedback. H.J.N. supervised the project.

## References

1. Dorsey, E. R. & Bloem, B. R. The Parkinson Pandemic-A Call to Action. JAMA Neurol 75, 9–10 (2018).

2. Nalls, M. A. et al. Identification of novel risk loci, causal insights, and heritable risk for Parkinson’s disease: a meta-analysis of genome-wide association studies. Lancet Neurol 18, 1091–1102 (2019).

3. Sherva, R. et al. Genome-wide association study of rate of cognitive decline in Alzheimer’s disease patients identifies novel genes and pathways. Alzheimers Dement 16, 1134–1145 (2020).

4. Iwaki, H. et al. Genetic risk of Parkinson disease and progression:: An analysis of 13 longitudinal cohorts. Neurol Genet 5, e348 (2019).

5. Maple-Grødem, J. et al. Association of GBA Genotype With Motor and Functional Decline in Patients With Newly Diagnosed Parkinson Disease. Neurology 96, e1036–e1044 (2021).

6. Ortega, R. A. et al. Association of Dual LRRK2 G2019S and GBA Variations With Parkinson Disease Progression. JAMA Netw Open 4, e215845 (2021).

7. Petrucci, S. et al. GBA-Related Parkinson’s Disease: Dissection of Genotype-Phenotype Correlates in a Large Italian Cohort. Mov Disord 35, 2106–2111 (2020).

8. Mahul-Mellier, A.-L. et al. The process of Lewy body formation, rather than simply α-synuclein fibrillization, is one of the major drivers of neurodegeneration. Proc Natl Acad Sci U S A 117, 4971–4982 (2020).

9. Braak, H. et al. Staging of brain pathology related to sporadic Parkinson’s disease. Neurobiol Aging 24, 197–211 (2003).

10. Braak, H., Rüb, U. & Del Tredici, K. Cognitive decline correlates with neuropathological stage in Parkinson’s disease. J Neurol Sci 248, 255–258 (2006).

11. Luk, K. C. et al. Pathological α-synuclein transmission initiates Parkinson-like neurodegeneration in nontransgenic mice. Science 338, 949–953 (2012).

12. Luk, K. C. et al. Intracerebral inoculation of pathological α-synuclein initiates a rapidly progressive neurodegenerative α-synucleinopathy in mice. J Exp Med 209, 975–986 (2012).

13. Luk, K. C. et al. Exogenous alpha-synuclein fibrils seed the formation of Lewy body-like intracellular inclusions in cultured cells. Proc Natl Acad Sci U S A 106, 20051–20056 (2009).

14. Polinski, N. K. et al. Best Practices for Generating and Using Alpha-Synuclein Pre-Formed Fibrils to Model Parkinson’s Disease in Rodents. J Parkinsons Dis 8, 303–322 (2018).

15. Volpicelli-Daley, L. A. et al. Exogenous α-synuclein fibrils induce Lewy body pathology leading to synaptic dysfunction and neuron death. Neuron 72, 57–71 (2011).

16. UniProt Consortium. UniProt: the universal protein knowledgebase in 2021. Nucleic Acids Res 49, D480–D489 (2021).

17. NCBI Resource Coordinators. Database resources of the National Center for Biotechnology Information. Nucleic Acids Res 46, D8–D13 (2018).

18. Alliance of Genome Resources Consortium. The Alliance of Genome Resources: Building a Modern Data Ecosystem for Model Organism Databases. Genetics 213, 1189–1196 (2019).

19. van Diepen, M. T. et al. The molluscan RING-finger protein L-TRIM is essential for neuronal outgrowth. Mol Cell Neurosci 29, 74–81 (2005).

20. Johnston, M. J. et al. High-resolution structural genomics reveals new therapeutic vulnerabilities in glioblastoma. Genome Res 29, 1211–1222 (2019).

21. Iwaki, H. et al. Genomewide association study of Parkinson’s disease clinical biomarkers in 12 longitudinal patients’ cohorts. Mov Disord 34, 1839–1850 (2019).

22. The National Genomics Research Library v5.1, Genomics England. 10.6084/m9.figshare.4530893/7. 2020.

23. GTEx Consortium. The GTEx Consortium atlas of genetic regulatory effects across human tissues. Science 369, 1318–1330 (2020).

24. Balastik, M. et al. Deficiency in ubiquitin ligase TRIM2 causes accumulation of neurofilament light chain and neurodegeneration. Proc Natl Acad Sci U S A 105, 12016– 12021 (2008).

25. Ylikallio, E. et al. Deficiency of the E3 ubiquitin ligase TRIM2 in early-onset axonal neuropathy. Hum Mol Genet 22, 2975–2983 (2013).

26. Goldman, J. E., Yen, S. H., Chiu, F. C. & Peress, N. S. Lewy bodies of Parkinson’s disease contain neurofilament antigens. Science 221, 1082–1084 (1983).

27. Trojanowski, J. Q. & Lee, V. M. Aggregation of neurofilament and alpha-synuclein proteins in Lewy bodies: implications for the pathogenesis of Parkinson disease and Lewy body dementia. Arch Neurol 55, 151–152 (1998).

28. Fares, M. B., Jagannath, S. & Lashuel, H. A. Reverse engineering Lewy bodies: how far have we come and how far can we go? Nat Rev Neurosci 22, 111–131 (2021).

29. Rocha, E. M. et al. Progressive decline of glucocerebrosidase in aging and Parkinson’s disease. Ann Clin Transl Neurol 2, 433–438 (2015).

30. Gan-Or, Z., Liong, C. & Alcalay, R. N. GBA-Associated Parkinson’s Disease and Other Synucleinopathies. Curr Neurol Neurosci Rep 18, 44 (2018).

31. Park, J.-S., Davis, R. L. & Sue, C. M. Mitochondrial Dysfunction in Parkinson’s Disease: New Mechanistic Insights and Therapeutic Perspectives. Curr Neurol Neurosci Rep 18, 21 (2018).

32. Buniello, A. et al. The NHGRI-EBI GWAS Catalog of published genome-wide association studies, targeted arrays and summary statistics 2019. Nucleic Acids Res 47, D1005–D1012 (2019).

33. Zheng, Q. et al. Dysregulation of Ubiquitin-Proteasome System in Neurodegenerative Diseases. Front Aging Neurosci 8, 303 (2016).

34. Sayers, E. W. et al. Database resources of the National Center for Biotechnology Information. Nucleic Acids Res 47, D23–D28 (2019).

35. Kang, X. et al. Tumor Necrosis Factor Inhibition and Parkinson Disease: A Mendelian Randomization Study. Neurology 96, e1672–e1679 (2021).

36. Calvo, S. E., Clauser, K. R. & Mootha, V. K. MitoCarta2.0: an updated inventory of mammalian mitochondrial proteins. Nucleic Acids Res 44, D1251–1257 (2016).

37. Blake, J. A. et al. Mouse Genome Database (MGD): Knowledgebase for mouse-human comparative biology. Nucleic Acids Res 49, D981–D987 (2021).

38. Gaare, J. J. et al. Rare genetic variation in mitochondrial pathways influences the risk for Parkinson’s disease. Mov Disord 33, 1591–1600 (2018).

39. Uhlén, M. et al. Proteomics. Tissue-based map of the human proteome. Science 347, 1260419 (2015).

40. Chen, P.-C. et al. Retinal Diseases and Parkinson Disease: A Population-Based Study. Front Neurosci 15, 679092 (2021).

41. Maetzler, W., Liepelt, I. & Berg, D. Progression of Parkinson’s disease in the clinical phase: potential markers. Lancet Neurol 8, 1158–1171 (2009).

42. Goetz, C. G. et al. Movement Disorder Society-sponsored revision of the Unified Parkinson’s Disease Rating Scale (MDS-UPDRS): scale presentation and clinimetric testing results. Mov Disord 23, 2129–2170 (2008).

43. McLaren, W. et al. The Ensembl Variant Effect Predictor. Genome Biol 17, 122 (2016).

44. Kolberg, L., Raudvere, U., Kuzmin, I., Vilo, J. & Peterson, H. gprofiler2 -- an R package for gene list functional enrichment analysis and namespace conversion toolset g:Profiler. F1000Res **9**, ELIXIR-709 (2020).

45. Durinck, S., Spellman, P. T., Birney, E. & Huber, W. Mapping identifiers for the integration of genomic datasets with the R/Bioconductor package biomaRt. Nat Protoc 4, 1184–1191 (2009).

46. Dobin, A. et al. STAR: ultrafast universal RNA-seq aligner. Bioinformatics 29, 15–21 (2013).

47. Li, H. et al. The Sequence Alignment/Map format and SAMtools. Bioinformatics 25, 2078– 2079 (2009).

48. Liao, Y., Smyth, G. K. & Shi, W. featureCounts: an efficient general purpose program for assigning sequence reads to genomic features. Bioinformatics 30, 923–930 (2014).

49. Law, C. W., Chen, Y., Shi, W. & Smyth, G. K. voom: Precision weights unlock linear model analysis tools for RNA-seq read counts. Genome Biol 15, R29 (2014).

50. Yu, G., Wang, L.-G., Han, Y. & He, Q.-Y. clusterProfiler: an R package for comparing biological themes among gene clusters. OMICS 16, 284–287 (2012).

51. Marek, K. et al. The Parkinson’s progression markers initiative (PPMI) - establishing a PD biomarker cohort. Ann Clin Transl Neurol 5, 1460–1477 (2018).

52. Plagnol, V., Smyth, D. J., Todd, J. A. & Clayton, D. G. Statistical independence of the colocalized association signals for type 1 diabetes and RPS26 gene expression on chromosome 12q13. Biostatistics 10, 327–334 (2009).

53. Witte, M. D. et al. Ultrasensitive in situ visualization of active glucocerebrosidase molecules. Nat Chem Biol 6, 907–913 (2010).

