## Supplementary Note for "Progression GWAS followed by functional characterization implicates E3 ubiquitin ligase *TRIM2* as a potential genetic modifier of Parkinson’s disease progression"

1. AMP-PD and PPMI GWAS
(1) PD progression rate distribution

PD progression rate was defined as the annual rate of change in MDS-UPDRS total score. The scores were normally distributed in both datasets (**Supplementary Figure 1**). An outlier with the rate of 60, derived from two time points, was excluded in the downstream analysis that used the PPMI dataset.

(2) Potential confounders: genetic relatedness, population structure, and other factors

To assess the genetic relatedness of individuals, pairwise identity-by-descent (IBD) estimate was calculated by averaging PLINK’s pi-hat on pruned variants (r^2^<0.2)^1^. Considering that first-degree relatives are expected to have IBD estimate of 0.5, second-degree relatives to have IBD of 0.25, and third-degree relatives to have IBD of 0.125, all the individuals in our PPMI dataset are genetically unrelated (IBD estimate < 0.1). Within the AMP-PD dataset, 19 individuals from relatedness pairs with PLINK’s pi-hat value > 0.125 were excluded from the analysis amounting to the final sample size of 1,114.

Population structure was evaluated using PLINK’s multi-dimensional scaling on pruned variants (r^2^<0.2). In the PPMI dataset, no clear genetic clustering was observed. After visual inspection of scatter plots with the first 6 components, the first 3 components were selected to be used as covariates in association analysis. There was no clear clustering of individuals by sex or clinical sites on the first two components of MDS. In the AMP-PD dataset, no clear genetic clustering was observed. After a visual inspection of the first 10 MDS components, the first 3 were selected to be used as covariates in the association tests. No clear clustering of individuals by sex or study was observed in the first two MDS components.

(3) GWAS quantile-quantile (Q-Q) plot assessment

The Q-Q plots of observed p-values and expected p-values largely follow the straight line in both datasets, with inflation factor close to 1 (PPMI λ_GC_ = 1.014, AMP-PD λ_GC_ = 1.005), indicating that genomic correction is not required (**Supplementary Figure 1c-d**).

(4) *TRIM2* locus conditional analysis

We applied two methods, GCTA-COJO and SuSiE, for the conditional analysis of *TRIM2* locus^2–4^. For CGTA-COJO analysis, the locus was defined by the following steps: (1) extend out to the most distant variants with r^2^>0.3 with lead SNP, (2) include SNPs with MAF>0.01 as recommended by GCTA-COJO authors. As a result, the tested locus included 334 SNPs in chr4:153,251,332-153,350,860 (~100kb). Conditional analysis was run for every variant in this locus. After conditioning on rs72729647, the p-value of rs62323742 was not significant (p=0.087; author recommended threshold P<1E-5), providing no evidence of a second distinct association in the *TRIM2* locus.

SuSiE determines the number of distinct signals in a region (default max 10) and returns posterior probabilities for each variant within each distinct signal. We ran SuSiE on individual data with and without the same covariates used in GWAS in the 1Mb region of *TRIM2* locus. Only one variant with high posterior probability was detected, suggesting that there is no second distinct association signal.

(5) Pathway analysis of the PFF-induced DEG and the three-way comparisons between PFF-induced DEG, AMP-PD, and PPMI GWAS genes with suggestive association

Pathway analyses on 3,123 PFF-induced DEGs with human orthologs identified 1,978 gene sets associated with α-syn aggregation **(Supplementary Table 6)**. Specifically, initial α-syn aggregate formation (D7 vs. PBS) was associated with transmembrane signaling receptor activity and G protein-coupled receptor (GPCR) activity. Maturation to LB-like aggregates (D21 vs. D14) was associated with extracellular matrix, calcium ion binding, integrin binding, GPCR, type I interferon, and inflammatory response. The overall α-syn aggregation process was associated with pathways such as GPCR signaling, neuronal system, neuroactive ligand-receptor interaction, learning/memory, and neuron projection.

(6) Pathway analysis of suggestive associations from AMP-PD and PPMI GWAS

For the AMP-PD GWAS, pathway analyses on the 453 genes from 161 genomic regions with suggestive associations (P < 1E-5, **Supplementary Table 2**) identified 19 Gene Ontology (GO) terms and 1 pathway, including cytokine and chemokine-related gene sets (adjusted-P < 0.05; **Supplementary Table 4**), suggesting a role for immune system dysfunction in PD progression. For the PPMI GWAS, pathway analyses on the 1,303 genes from 162 genomic regions with suggestive associations (P < 1E-5, **Supplementary Table 3**) identified 4 GO terms, including purinergic receptor and endopeptidase inhibitor activities (**Supplementary Table 4**).

(7) Variant aggregate analysis (SKAT-O)

The Q-Q plot of observed p-values and expected p-values from the four SKAT-O analyses largely follow the straight line, with inflation factor close to 1 (**Supplementary Figure 2a**).

As a targeted analysis, two genes were investigated. *GBA*, a gene whose cumulative non-synonymous mutations have been associated with PD progression in other studies via targeted approaches, and *TRIM2*, our candidate gene^5^. *GBA* association with PD progression did not replicate in our analysis (best SKAT-O p=0.235, **Supplementary Figure 2b**, **Supplementary Table 2**). Of the four tests conducted for *TRIM2*, two tests generated results, as no LoF mutations were detected in our dataset. The resulting test suggests an aggregate effect of missense mutations in slower PD progression rate (**Supplementary Figure 2c**, **Supplementary Table 5**).

2. GWAS results prioritization
(1) Interpreting PFF-induced DEGs: Recapitulation of LB formation process

Mahul-Mellier *et al.* observed alpha-synuclein pS129+ neuritic inclusions 4 days after PFF treatment^6^. On Day 7, they observed filamentous alpha-synuclein aggregates in dendrites and some axons, with few aggregates in the cell body. The number of filamentous aggregates in the neurites and cell bodies increased on Day 14. On Day 21, filamentous, ribbon-like, and round Lewy Body-like inclusions were observed.

(2) GWAS genes with suggestive associations and PFF-induced DEGs comparison

To identify candidate genes and pathways underlying PD progression, we compared 453 AMP-PD GWAS genes with suggestive associations (P < 1E-5), 1,303 PPMI GWAS genes with suggestive associations (P < 1E-5), and 3,123 PFF-induced DEGs. We found that RNA binding fox-1 homolog 1 (*RBFOX1*), a gene implicated in neurodevelopmental disorders, was shared across all gene sets (**Figure 2b**, **Supplementary Table 7)**.

Pairwise comparisons detected additional candidates, including those previously implicated in PD (**Figure 2b**, **Supplementary Table 7**). (1) AMP-PD vs. PPMI GWAS genes with suggestive associations: 8 genes were shared, including polypeptide N-acetylgalactosaminyltransferase 13 (*GALNT13*), MIR4435-2 host gene (*MIR4435-2HG*), parkin RBR E3 ubiquitin protein ligase (*PRKN*), tRNA splicing endonuclease subunit 15 (*TSEN15*), a novel gene annotated with TSEN15 antisense transcript, long intergenic non-protein coding RNA 1445 (*LINC01445*), and two Y_RNA associated genes. (2) AMP-PD GWAS genes with suggestive associations vs. PFF-induced DEGs: 34 genes were shared, including synuclein alpha (*SNCA)*, semaphorin 3E (*SEMA3E*) and GPRIN family member 3 (*GPRIN3*). 71% (24) and 91% (31) were present in the pathways enriched in AMP-PD GWAS genes with suggestive associations and PFF-induced DEGs, respectively, suggesting that these genes represent key pathways detected in each PD progression analysis. (3) PPMI GWAS genes with suggestive associations vs. PFF-induced DEGs: 94 genes were shared, including rogdi atypical leucine zipper (*ROGDI*), tripartite motif containing 36 (*TRIM36*), as well as *TRIM2*, the top candidate from our PPMI PD progression GWAS. 3% (3) and 95% (89) were present in the pathways enriched in PPMI GWAS genes with suggestive associations and PFF-induced DEGs, respectively. The low number of genes captured in PPMI pathway comparison is likely driven by only a few pathways detected in PPMI GWAS.

At the pathway-level, GPCR activities were shared across all three gene sets (**Supplementary Table 4, Supplementary Table 6)**. Cell adhesion, plasma membrane, calcium ion binding, and cytokine receptors were commonly detected in AMP-PD GWAS genes with suggestive associations and PFF-induced DEGs. Endopeptidase inhibition was commonly detected in PPMI GWAS genes with suggestive associations and PFF-induced DEGs.

(3) Colocalization and Brain expression Quantitative Trait Loci (eQTL)

To investigate statistical causality of GWAS hits, we performed a colocalization analysis between PD-progression GWAS loci and eQTLs mapped by GTEx in 13 different brain regions. We examined 245 AMP-PD loci and 249 PPMI loci within a cis window (1Mb up/downstream) of each GWAS hit. Out of 176 million GTEx brain eQTLs, we probed 52,299 eQTLs that have a common position with PD progression GWAS loci. We calculated the posterior probability of GWAS and eQTL signals to share the same location and identified no GWAS loci with posterior probability of colocalization greater than 0.5.

(4) GTEx brain eQTLs in *TRIM2* locus

We interrogated our two SNPs in the GTEx’s brain eQTL data (13 different brain regions) to evaluate whether these SNPs are correlated with any gene expressions. We evaluated all 4 genes within 500kb window of our two SNPs and found that only TRIM2 expression level was associated with these two SNPs. Specifically, rs72729647 A allele was correlated with increased expression of TRIM2 in brain cortex (P=5.35E-3) and brain spinal cord (P=3.95E-3). rs62323742 G allele was correlated with decrease in TRIM2 expression in brain cortex (P=3.64E-3) and increase in brain spinal cord (P=1E-3). The normalized TRIM2 expression levels by genotype are shown in **Supplementary Figure 3**.

(5) Brain enhancers and TADs for TRIM2 GWAS variants

For physical interaction assessment of TRIM2 genomic locus, 5kb DNA contact maps and 3D genome architecture maps were obtained from a public in-situ Hi-C data in GB_523 cell line, generated using glioblastoma stem cells (GSCs) from patient diagnosed with glioblastoma (GBM). The topological associated domain (TAD) and interaction maps were generated using Dixon et. al pipeline^7^. Chromatin loops were predicted by Peakachu^8^. 3D Genome Browser was used for visualization and to generate the plots^9^. To this end, we found that the TRIM2 GWAS SNP locus and TRIM2 are within the same TAD with contacts with promoter of TRIM2, suggesting a regulatory role (**Figure 3b**).

For enhancer activity assessment, we looked at Nott et.al datasets profiling chromatin and promoter activity in cell nuclei isolated from human brains^10^. The cell types profiled were microglia, neurons, oligodendrocytes, and astrocytes. Nuclei were subjected to ATAC-seq and H3K27Ac and H3K4me3 ChIP-seq. While the two GWAS variants did not directly overlap with any regulatory signals in the locus, multiple linked variants did overlap with open chromatin marks. A low-frequency (1000 Genomes AF = 0.04) TRIM2 intronic variant sharing the LD with the top SNP (rs72729639, 1000 Genomes EUR D’=0.96, N=503) overlaps with an open chromatin peak, which is also a potential CTCF site (ENCODE datasets^7^). Another linked TRIM2 intronic variant (rs13117024, 1000 Genomes AF = 0.27, 1000 Genomes EUR D’=0.93, N=503) overlaps with open chromatin peak only in neurons^10^ (**Supplementary Figure 4**).

(6) *TRIM2* GWAS SNPs in Iwaki and GeL datasets

We interrogated our two SNPs in the Iwaki dataset, which contains summary statistics of GWAS meta-analysis on 25 phenotypes from 12 longitudinal cohorts^11^. The two SNPs showed nominal associations with UPDRS3_scaled (motor examination; rs72729647 P=0.009; rs62323742 P=0.006) and time to motor fluctuation (rs62323742, P=0.01). In addition, rs72729647 showed weak association with UPDRS4_scaled (motor complication; P=0.06). The number of individuals and datasets included for each meta-analysis is shown in **Table 2**.

In the GeL dataset, both SNPs showed higher carrier frequencies in PD fast progressors compared to PD not-fast progressors (rs72729647, FP=0.41, NFP=0.33, Fisher’s exact P=0.29; rs62323742, FP=0.62, NFP=0.55, Fisher’s exact P=0.11; **Table 2**).

3. Additional genetic candidates for potential follow-up with high-throughput functional screening

Intersection of the two sets of GWAS genes with suggestive associations and PFF-DEGs identified 137 genetic candidates for PD progression that are supported by at least two datasets. These candidates include known familial PD genes (*PRKN* and *SNCA),* and several genes that are relevant to PD cellular pathophysiology. Notably, RBFOX1 is elevated in PD patients’ iPSC-derived dopaminergic neurons, dysregulating RNA splicing and impacting mitochondrial and neuron activity-related genes^12^. *RBFOX1* is also associated with neurofibrillary tangles^13^. RBFOX1 binds to ataxin-2, which causes neurodegenerative spinocerebellar ataxia type 2, when mutated^14^. *TRIM36*, which is in the same family as our top candidate *TRIM2*, is downregulated in substantia nigra pars compacta of PD patients^15^. *ROGDI* mutations cause Kohlschutter-tonz syndrome, a neurodegenerative disorder with progressive dementia (OMIM: 226750). SEMA3E and GPRIN3 play a key role in neurodevelopment and neuronal excitability in dopaminergic neurons, respectively^16,17^. Taken together, our time-series genomic and transcriptomic analyses of PD progression identified several genes with established roles in motor dysfunction and PD cellular physiology, as well as novel candidates with heretofore unknown links to PD.

These candidate genes, combined with our shared pathway results, suggest that neuroinflammation, endopeptidase inhibition, cell adhesion, calcium signaling, GPCR signaling, and ubiquitination may be critical drivers of PD progression. Purinergic receptors are also of interest, considering its potential role in neurodegeneration with motor impairments via mitochondrial dysfunction, although it was identified only once in our PPMI GWAS^18,19^.

4. Peripheral biomolecule analyses - Results
(1) Omics analyses of peripheral biofluids detect molecular changes associated with PD severity.

To identify molecular changes associated with PD severity, we analyzed the whole blood, CSF, and plasma from the PPMI cohort. Several analyses were performed based on the availability of patient data: (a) RNA-seq analysis in whole blood from 420 patients to identify gene expression changes associated with total MDS-UPDRS (**Supplementary Table 9**); (b) DNA-methylation analysis in whole blood from 297 patients to identify epigenetic changes associated with total MDS-UPDRS (**Supplementary Table 10**); and (c) proteomic analysis from 117 patients to identify differentially abundant proteins in matched CSF and plasma (**Supplementary Table 11**). In association with PD severity, we identified 1201 (594 ↑; 607 ↓) differentially expressed genes and 1865 CpGs (1019 hypermethylated; 846 hypomethylated) corresponding to 1453 differentially methylated genes in whole blood. Additionally, 162 proteins in the CSF and 293 proteins in the plasma were found to be differentially abundant with PD severity.

To explore peripheral biomarker candidates of PD severity, we compared genes that were differentially expressed to those that were differentially methylated in whole blood and to the proteins differentially abundant in CSF or plasma. Of these, 65 genes were found to be both differentially expressed and methylated. As expected, the methylation states and expression level for these genes were negatively correlated (Spearman ρ = -0.3; P = 0.0096), suggesting that the hypermethylated CpGs lead to transcript inactivation while the hypomethylated CpGs lead to transcript activation of these genes (**Supplementary Table 12)**. Of those, 19 had promoter-specific methylation, corroborating that transcription of these genes are dysregulated by the detected CpGs (**Supplementary Table 12**)^20,21^. Additionally, we identified 8 plasma proteins and 11 CSF proteins overlapping with differentially expressed genes in whole blood; and 13 plasma proteins and 4 CSF proteins overlapping with differentially methylated genes in whole blood, suggesting that methylation and expression changes of these genes may have translated to differences in the protein levels (**Supplementary Table 12**).

(2) PD *cis*-eQTL and genetic overlap prioritize PD severity-associated molecular changes.

To prioritize peripheral biomarker candidates based on genetic support, we interrogated them in the 15,401 genes identified from our re-analysis of PPMI PD blood *cis*-eQTLs (adjusted P < 0.05). We found 47 genes that were both differentially expressed and methylated to be mapped to PD cis-eQTLs, including leucine-rich repeat kinase 2 (*LRRK2*), proteasome subunit beta type-8 (*PSMB8*), and caspase recruitment domain family member 11 (*CARD11*) (**Supplementary Table 13**). Separately, we compared 1,747 GWAS ‘suggestive and significant association’ (SSA, P < 1E-5) genes with those differentially expressed or methylated in the periphery and identified four genes that were common across all three sets: *CARD11*, *MED13L,* spectrin alpha non-erythrocytic 1 (*SPTAN1*), and family with sequence similarity 124 member B (*FAM124B*). An additional 39 differentially expressed genes and 44 differentially methylated genes were found to be common with GWAS SSA genes, providing more PD-severity biomolecules supported by genetic evidence (**Supplementary Table 14**). In summary, the overlap of several genes that were both differentially expressed and methylated, with PD cis-eQTLs and PD GWAS genes suggests that these genes, which include *CARD11, MED13L, SPTAN1*, and *FAM124B*, may be good candidates to further test their utility as peripheral biomarkers of PD progression.

(3) Implications for future peripheral biomarker discovery

Our top candidates with known CNS functions arise from 3 categories: (1) Molecular changes supported by regulatory mechanism – 72% (47/65) of our differentially expressed and methylated genes were mapped to PD blood eQTLs, providing potential gene expression mechanisms regulated by specific CpGs and regulatory variants. These include *LRRK2*, a causal gene for familial PD and a predominant risk factor for idiopathic PD when mutated, *CARD11*, a gene associated with multiple sclerosis, and *PSMB8*, a gene upregulated in PD-rat models^20–24^. (2) Molecular changes supported by GWAS – our 4 differentially expressed and methylated genes were supported by GWAS, identifying SNPs potentially impacting clinical PD progression via peripheral gene expression and methylation changes. All four are involved in neurological disorders: *CARD11* (see above)*, MED13L,* whose haploinsufficiency causes a movement impairment syndrome including hypotonia and ataxia*, SPTAN1*, a gene associated with schizophrenia*,* and *FAM124B,* a gene associated with neurodevelopmental disorders^25,26^. Of note, *MED13L* is one of our four GWAS SA / PFF-induced DEGs. Thus, we propose *MED13L* as a candidate for both regulator and peripheral biomarker of disease progression. (3) Protein changes in both biofluids – Our top candidates include those repeatedly associated, including NF-L (blood for PD severity and progression, CSF and plasma for PD risk), NTproBNP (blood for PD motor and cognitive decline) and GDF15 (serum for PD and age-of-onset of Parkinsonism)^27–31^.

5. Peripheral biomolecule analyses - Methods
(1) Whole blood RNA-seq data from the PPMI cohort

Raw FASTQ files of whole blood RNA-seq samples from the PPMI PD cohort were obtained from the PPMI LONI data repository. A total of 1,211 samples from 420 PD patients across four time points, including baseline and Years 1-3, were used for the RNA-seq analysis. Raw paired-end reads were first trimmed using Trimmomatic version 0.36 and aligned to version GRCh38 of the human genome using STAR version 2.5^32,33^. Read counts were then generated using featureCounts from the Subread package version 1.6^34^. MultiQC was used to compile and to generate the summary statistics of the results per sample for QC evaluation^35^.

The feature counts were exported to R (4.0.2) and filtered for low expression using a CPM (count per million) threshold of 1 in at least 400 samples. Principal component analysis (PCA) was conducted on the filtered count matrix, revealing no distinct clusters by visit, gender or MDS-UPDRS score. The raw counts were normalized using the TMM normalization and the *voom* function from the *limma* package was applied to incorporate precision weights accounting for the mean-variance relationship and minimize heteroscedasticity^36^. A linear model was fit on the voom-normalized data adjusting for gender, with total MDS-UPDRS score as the variable of interest. In addition, due to the longitudinal nature of the study, the patient ID was used as a blocking variable to adjust for individual-specific variability in gene expression data and the *duplicatecorrelation* function was used to calculate the intrablock correlation across all genes^37^. Differential gene expression associated with a unit change in the total MDS-UPDRS score was calculated using an empirical Bayes statistic in limma. The raw p-values were adjusted for multiple comparisons using the Benjamini-Hochberg correction and the adjusted p-value of 0.05 was used to characterize the statistically significant genes.

(2) PPMI DNA methylome

Whole Blood DNA methylation data (Illumina 850K Infinium MethylationEPIC BeadChip) from 315 PD patients was obtained via the PPMI data LONI repository (project 140). A total of 1,243 samples for at least 3 study visits over an approximately 3-year period were included in the analyses. Clinical data, including MDS-UPDRS scores, were downloaded from the PPMI LONI data repository.

Sample barcodes were used to infer the plate randomization design revealing that samples from the same patient were plated together in the same row, unless there were technical replicates^38^. Samples with ≥1% of CpG sites with a detection p-value > 0.05 and samples with log2 (median methylated intensity) + log2 (median unmethylated intensity) < 21 were filtered out. Sex was predicted from X and Y chromosome intensities using the *minfi* *getSex* function and 4 samples whose reported sex did not match their predicted sex were excluded^39^. Sample identity was verified using the 59 single nucleotide polymorphism (SNP) probes included on the array and 3 mismatches were removed. After QC, the final dataset for analysis comprised of 950 samples from 297 patients.

Linear models were fitted on methylation M values for the total MDS-UPDRS score using *limma* and included a random effect for patient ID to account for the longitudinal repeated measures^37^. The final model to test for differentially methylated CpG loci included estimated cell-type proportions, plate-effects, sex, and race as covariates. Probes with FDR-adjusted p-values < 0.05 were classified as differentially methylated probes (DMPs). Using the Illumina manifest, we annotated DMPs with their location relative to genes and CpG islands. Chromatin state was predicted using ChromHMM predictions in ENCODE for the GM12878 (lymphoblastic) cell line^40^. Genes annotated with DMPs were deemed as differentially methylated genes (DMGs) and were analyzed for pathway overrepresentation using GO, KEGG and Reactome databases using *gprofiler2* R package^41,42^.

(3) PD *cis*-eQTL analysis

The PPMI cis-eQTL was called from RNA-seq samples across all time points. Among all samples from a time point, the gene expression levels were first quantified as TPM and then inverse normalized to be a standard normal across samples. The PEER factors were calculated for each time point separately to account for batch effects. Across the genotypes of all samples, the genetic principal components were calculated to account for population stratification. For the eQTL calling, we pooled samples from all time points together and to handle the overlapped samples across time points, we considered a linear mixed effects model $y_{it}=C_{it}\alpha+X_{i}\beta+z_{i}+\epsilon_{it}$ where $i$ and $t$ index individuals and time points with $y$, $C$, and $X$ representing expression level, covariates, and genotypes separately. $z_{i}$ is an individual-level random effect term representing an individual-specific baseline expression level and $\epsilon_{it}$ is the residual noise. To solve this model exactly requires a huge computational burden so, instead, we took a two-stage approach to solve it approximately. In the first stage, we fit a null model ($\beta=0$) to obtain estimates of the variance $V_{r}$ and $V_{e}$ for random effects $z_{i}$ and $\epsilon_{it}$. In the second stage, we fit a generalized linear model $y_{it}=C_{it}\alpha+X_{i}\beta+e_{it}, e_{it}\sim N(0,\Sigma)$ with $\Sigma_{(it),(jt')}=V_{r}+V_{e}$ when $i=j$ and $t=t'$, $\Sigma_{(it),(jt')}=V_{r}$ when $i=j$ and $t\neq t'$, and $\Sigma_{(it),(jt')}=0$ when $i\neq j$. PEER factors and genetics principal components along with age and sex were included as covariates in eQTL calling. For each gene, the size of the cis-window being considered was 1Mb and all variants with minor allele frequency greater than 0.01 within the cis-window were tested for cis-eQTL.

**Supplementary References**

1.	Purcell, S. *et al.* PLINK: A Tool Set for Whole-Genome Association and Population-Based Linkage Analyses. *Am J Hum Genet* **81**, 559–575 (2007).

2.	Wang, G., Sarkar, A., Carbonetto, P. & Stephens, M. A simple new approach to variable selection in regression, with application to genetic fine mapping. *J R Stat Soc Series B Stat Methodol* **82**, 1273–1300 (2020).

3.	GCTA: A Tool for Genome-wide Complex Trait Analysis - PMC. https://www.ncbi.nlm.nih.gov/pmc/articles/PMC3014363/.

4.	Yang, J. *et al.* Conditional and joint multiple-SNP analysis of GWAS summary statistics identifies additional variants influencing complex traits. *Nat Genet* **44**, 369–375, S1-3 (2012).

5.	Gan-Or, Z., Liong, C. & Alcalay, R. N. GBA-Associated Parkinson’s Disease and Other Synucleinopathies. *Curr Neurol Neurosci Rep* **18**, 44 (2018).

6.	Mahul-Mellier, A.-L. *et al.* The process of Lewy body formation, rather than simply α-synuclein fibrillization, is one of the major drivers of neurodegeneration. *Proc Natl Acad Sci U S A* **117**, 4971–4982 (2020).

7.	Johnston, M. J. *et al.* High-resolution structural genomics reveals new therapeutic vulnerabilities in glioblastoma. *Genome Res* **29**, 1211–1222 (2019).

8.	Salameh, T. J. *et al.* A supervised learning framework for chromatin loop detection in genome-wide contact maps. *Nat Commun* **11**, 3428 (2020).

9.	Wang, Y. *et al.* The 3D Genome Browser: a web-based browser for visualizing 3D genome organization and long-range chromatin interactions. *Genome Biol* **19**, 151 (2018).

10.	Nott, A. *et al.* Brain cell type-specific enhancer-promoter interactome maps and disease-risk association. *Science* **366**, 1134–1139 (2019).

11.	Iwaki, H. *et al.* Genetic risk of Parkinson disease and progression:: An analysis of 13 longitudinal cohorts. *Neurol Genet* **5**, e348 (2019).

12.	Lin, L. *et al.* Molecular Features Underlying Neurodegeneration Identified through In Vitro Modeling of Genetically Diverse Parkinson’s Disease Patients. *Cell Rep* **15**, 2411–2426 (2016).

13.	Buniello, A. *et al.* The NHGRI-EBI GWAS Catalog of published genome-wide association studies, targeted arrays and summary statistics 2019. *Nucleic Acids Res* **47**, D1005–D1012 (2019).

14.	Sayers, E. W. *et al.* Database resources of the National Center for Biotechnology Information. *Nucleic Acids Res* **47**, D23–D28 (2019).

15.	Grünblatt, E. *et al.* Gene expression profiling of parkinsonian substantia nigra pars compacta; alterations in ubiquitin-proteasome, heat shock protein, iron and oxidative stress regulated proteins, cell adhesion/cellular matrix and vesicle trafficking genes. *J Neural Transm (Vienna)* **111**, 1543–1573 (2004).

16.	Ding, J. B., Oh, W.-J., Sabatini, B. L. & Gu, C. Semaphorin 3E-Plexin-D1 signaling controls pathway-specific synapse formation in the striatum. *Nat Neurosci* **15**, 215–223 (2011).

17.	Karadurmus, D. *et al.* GPRIN3 Controls Neuronal Excitability, Morphology, and Striatal-Dependent Behaviors in the Indirect Pathway of the Striatum. *J Neurosci* **39**, 7513–7528 (2019).

18.	Oliveira-Giacomelli, Á. *et al.* Purinergic Receptors in Neurological Diseases With Motor Symptoms: Targets for Therapy. *Front Pharmacol* **9**, 325 (2018).

19.	Tóth, A., Antal, Z., Bereczki, D. & Sperlágh, B. Purinergic Signalling in Parkinson’s Disease: A Multi-target System to Combat Neurodegeneration. *Neurochem Res* **44**, 2413–2422 (2019).

20.	Rui, Q., Ni, H., Li, D., Gao, R. & Chen, G. The Role of LRRK2 in Neurodegeneration of Parkinson Disease. *Curr Neuropharmacol* **16**, 1348–1357 (2018).

21.	Maltby, V. E. *et al.* Genome-wide DNA methylation changes in CD19+ B cells from relapsing-remitting multiple sclerosis patients. *Sci Rep* **8**, 17418 (2018).

22.	International Multiple Sclerosis Genetics Consortium (IMSGC) *et al.* Analysis of immune-related loci identifies 48 new susceptibility variants for multiple sclerosis. *Nat Genet* **45**, 1353–1360 (2013).

23.	Ogino, M. *et al.* Roles of PTEN with DNA Repair in Parkinson’s Disease. *Int J Mol Sci* **17**, 954 (2016).

24.	Sun, C., Jia, G., Wang, X., Wang, Y. & Liu, Y. Immunoproteasome is up-regulated in rotenone-induced Parkinson’s disease rat model. *Neurosci Lett* **738**, 135360 (2020).

25.	Ranganathan, M. *et al.* Analysis of circulating exosomes reveals a peripheral signature of astrocytic pathology in schizophrenia. *World J Biol Psychiatry* **23**, 33–45 (2022).

26.	Batsukh, T. *et al.* Identification and characterization of FAM124B as a novel component of a CHD7 and CHD8 containing complex. *PLoS One* **7**, e52640 (2012).

27.	Lin, C.-H. *et al.* Blood NfL: A biomarker for disease severity and progression in Parkinson disease. *Neurology* **93**, e1104–e1111 (2019).

28.	Oosterveld, L. P. *et al.* CSF or serum neurofilament light added to α-Synuclein panel discriminates Parkinson’s from controls. *Mov Disord* **35**, 288–295 (2020).

29.	Choe, C.-U. *et al.* Subclinical Cardiac Microdamage, Motor Severity, and Cognition in Parkinson’s Disease. *Mov Disord* **35**, 1863–1868 (2020).

30.	Maetzler, W. *et al.* GDF15/MIC1 and MMP9 Cerebrospinal Fluid Levels in Parkinson’s Disease and Lewy Body Dementia. *PLoS One* **11**, e0149349 (2016).

31.	Yao, X. *et al.* Serum Growth Differentiation Factor 15 in Parkinson Disease. *Neurodegener Dis* **17**, 251–260 (2017).

32.	Dobin, A. *et al.* STAR: ultrafast universal RNA-seq aligner. *Bioinformatics* **29**, 15–21 (2013).

33.	Bolger, A. M., Lohse, M. & Usadel, B. Trimmomatic: a flexible trimmer for Illumina sequence data. *Bioinformatics* **30**, 2114–2120 (2014).

34.	Liao, Y., Smyth, G. K. & Shi, W. featureCounts: an efficient general purpose program for assigning sequence reads to genomic features. *Bioinformatics* **30**, 923–930 (2014).

35.	Ewels, P., Magnusson, M., Lundin, S. & Käller, M. MultiQC: summarize analysis results for multiple tools and samples in a single report. *Bioinformatics* **32**, 3047–3048 (2016).

36.	Law, C. W., Chen, Y., Shi, W. & Smyth, G. K. voom: Precision weights unlock linear model analysis tools for RNA-seq read counts. *Genome Biol* **15**, R29 (2014).

37.	Ritchie, M. E. *et al.* limma powers differential expression analyses for RNA-sequencing and microarray studies. *Nucleic Acids Res* **43**, e47 (2015).

38.	Marek, K. *et al.* The Parkinson’s progression markers initiative (PPMI) - establishing a PD biomarker cohort. *Ann Clin Transl Neurol* **5**, 1460–1477 (2018).

39.	Aryee, M. J. *et al.* Minfi: a flexible and comprehensive Bioconductor package for the analysis of Infinium DNA methylation microarrays. *Bioinformatics* **30**, 1363–1369 (2014).

40.	Ernst, J. & Kellis, M. Chromatin-state discovery and genome annotation with ChromHMM. *Nat Protoc* **12**, 2478–2492 (2017).

41.	Kolberg, L., Raudvere, U., Kuzmin, I., Vilo, J. & Peterson, H. gprofiler2 -- an R package for gene list functional enrichment analysis and namespace conversion toolset g:Profiler. *F1000Res* **9**, ELIXIR-709 (2020).

42.	Phipson, B., Maksimovic, J. & Oshlack, A. missMethyl: an R package for analyzing data from Illumina’s HumanMethylation450 platform. *Bioinformatics* **32**, 286–288 (2016).
