## Supplementary Figures for "Progression GWAS followed by functional characterization implicates E3 ubiquitin ligase *TRIM2* as a potential genetic modifier of Parkinson’s disease progression"

**Supplementary Fig 1. AMP-PD and PPMI phenotype and association statistics distribution. a-b.** Distribution of annual rate of change in MDS-UPDRS total score in AMP-PD (a) and PPMI (b). **c-d.** Q-Q plot in AMP-PD (c) and PPMI (d).

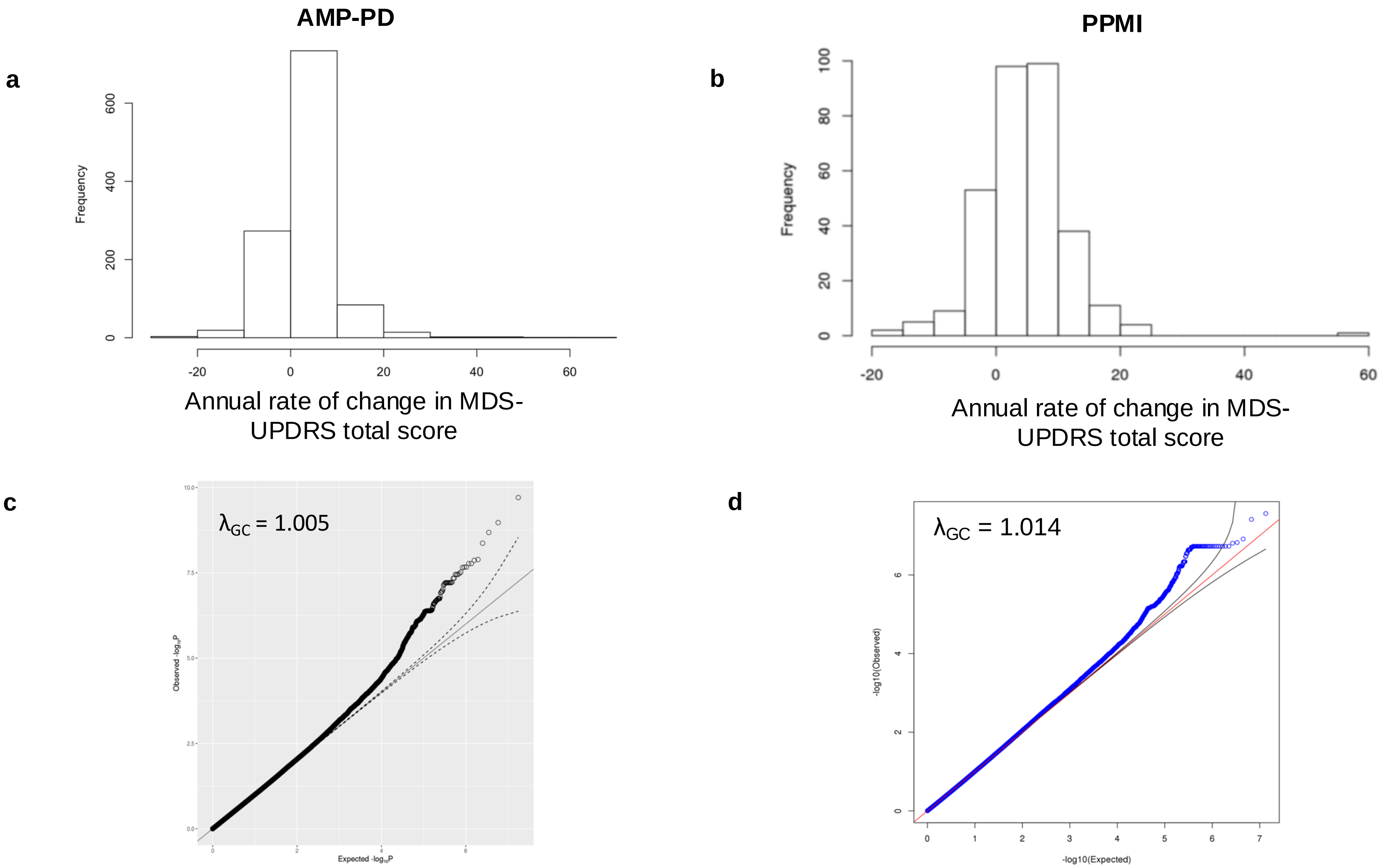

**Supplementary Fig 2. PD progression rate variant aggregate test. a.** Q-Q- plots by variant types and rare variant weights. **b-c.** PD progression rate of missense carriers stratified by cumulative allele count in GBA (c) and in TRIM2 (d).

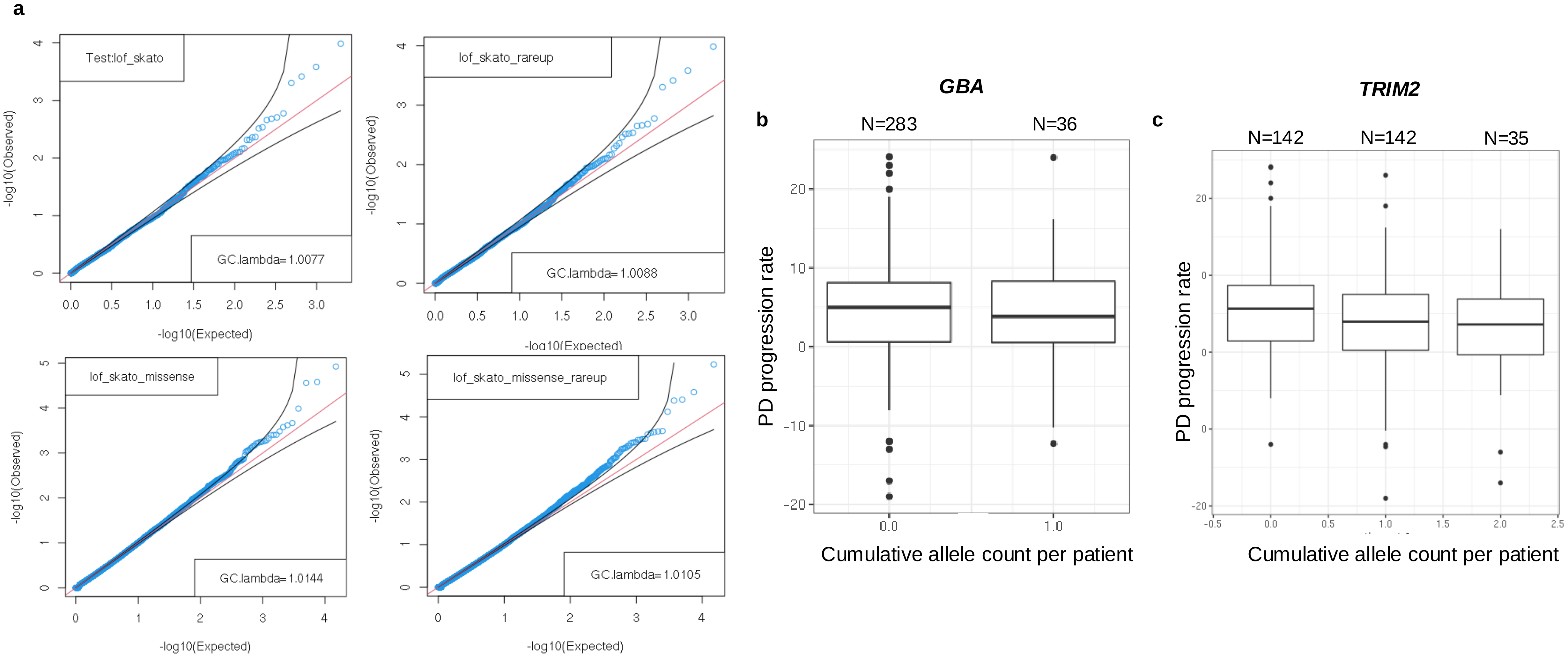

**Supplementary Fig 3. Brain eQTL evidence for TRIM2 GWAS variants. a-b.** rs72729647 (a) and rs62323742 (b) are correlated with TRIM2 transcript expression level in brain tissues.

**a**

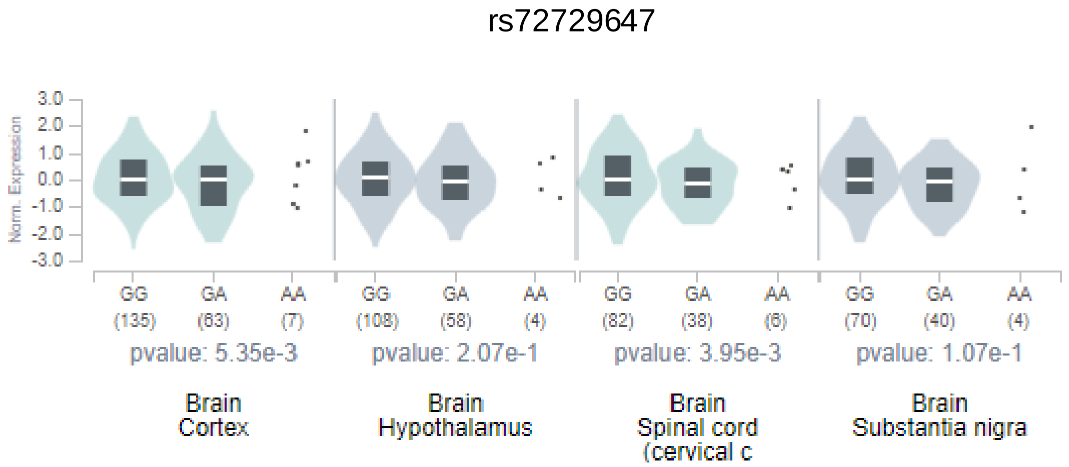


**b**

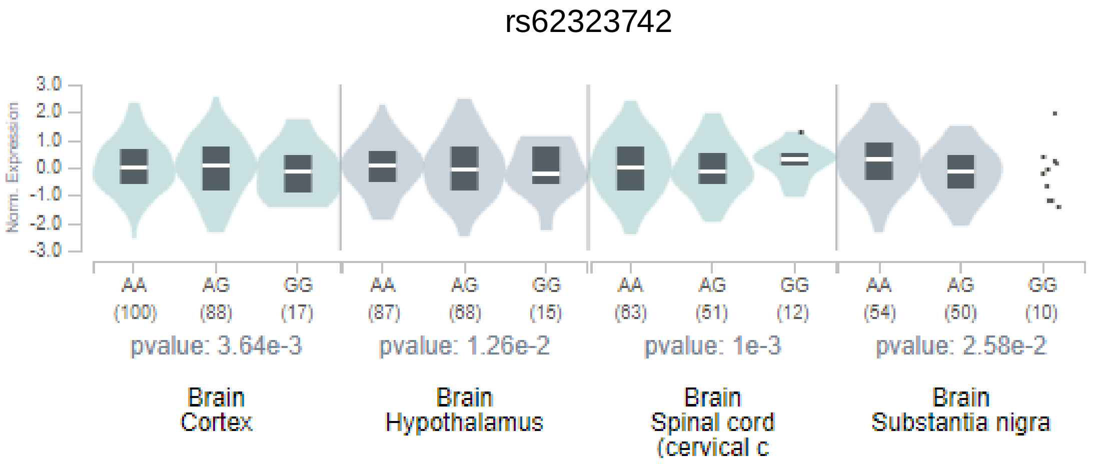

**Supplementary Fig 4. Brain enhancers in TRIM2 GWAS SNP locus.** The TRIM2 GWAS locus overlaps with brain enhancer signals. **a.** Two variants (rs72729639, rs13117024) that are linked to the TRIM2 GWAS SNPs (rs62323742, rs72729647) directly overlap with ATAC-seq signals in microglia, neurons, oligodendrocyte and astrocytes (Nott *et al.* dataset). **b.** rs72729639 specifically overlaps with CTCF peak identified in glioblastoma cell line (ENCODE dataset).

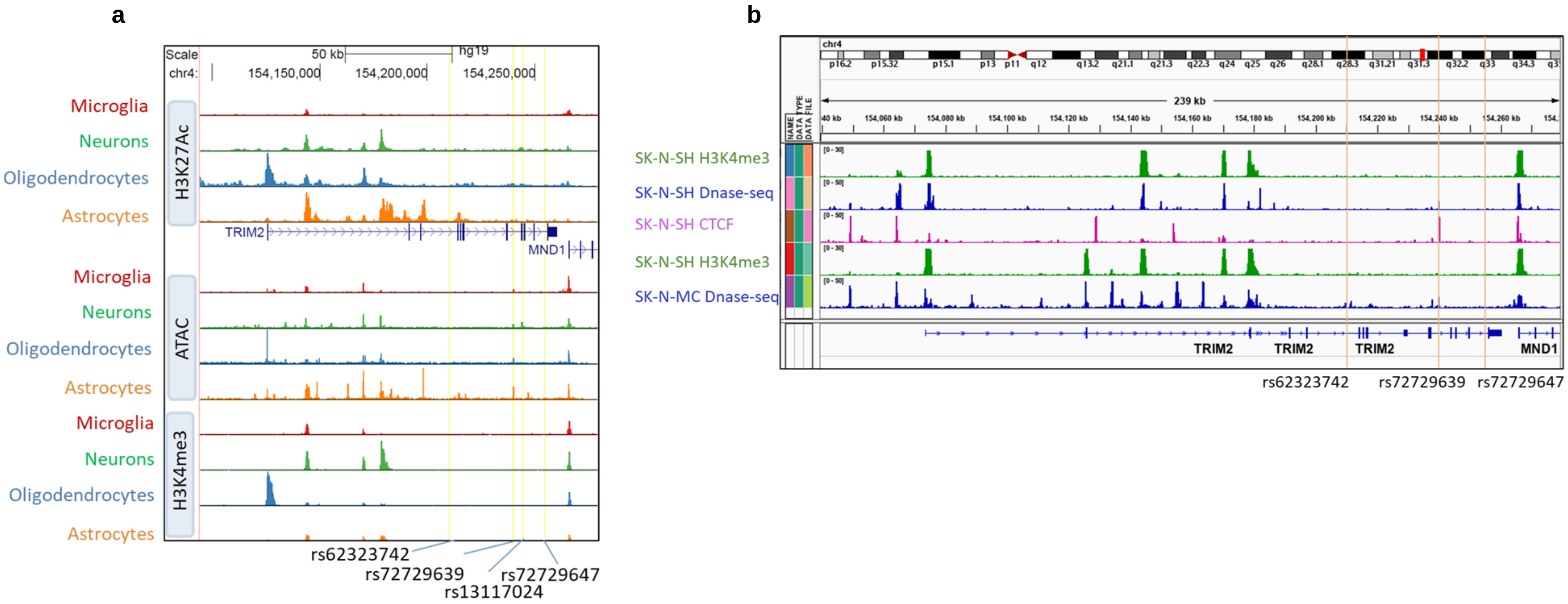

**Supplementary Fig 5. TRIM2 regulates neurofilament-L protein level in primary neurons. a.** Representative immunoblot images of CD1 neuronal lysates treated with 1 µM non-targeting control (NT Ctrl) or murine TRIM2 (mTRIM2) targeting Accell siRNAs and transduction with control (LacZ Ctrl) or human full-length TRIM2 (hTRIM2) lentivirus (LV) for 14d. **b-c.** Quantification of Trim2 mRNA (b) and TRIM2 protein (c) levels relative to control in a. ****p<0.0001 by unpaired Student's t-test; n=3/group. **d.** Quantification of TRIM2 overexpression level in neurons transduced with hTRIM2 LV compared to control. ***p=0.0001 by unpaire d Student's t-test; n=3/group. **e-f.** Quantification of neurofilament (NF-L) protein level in neurons treated with mTRIM2 siRNAs (e) or transduced with hTRIM2 LV (f). **p=0.0013 *p=0.0415 by unpaired Student's t-test; n=3/group. Error bars represent mean ± SEM.

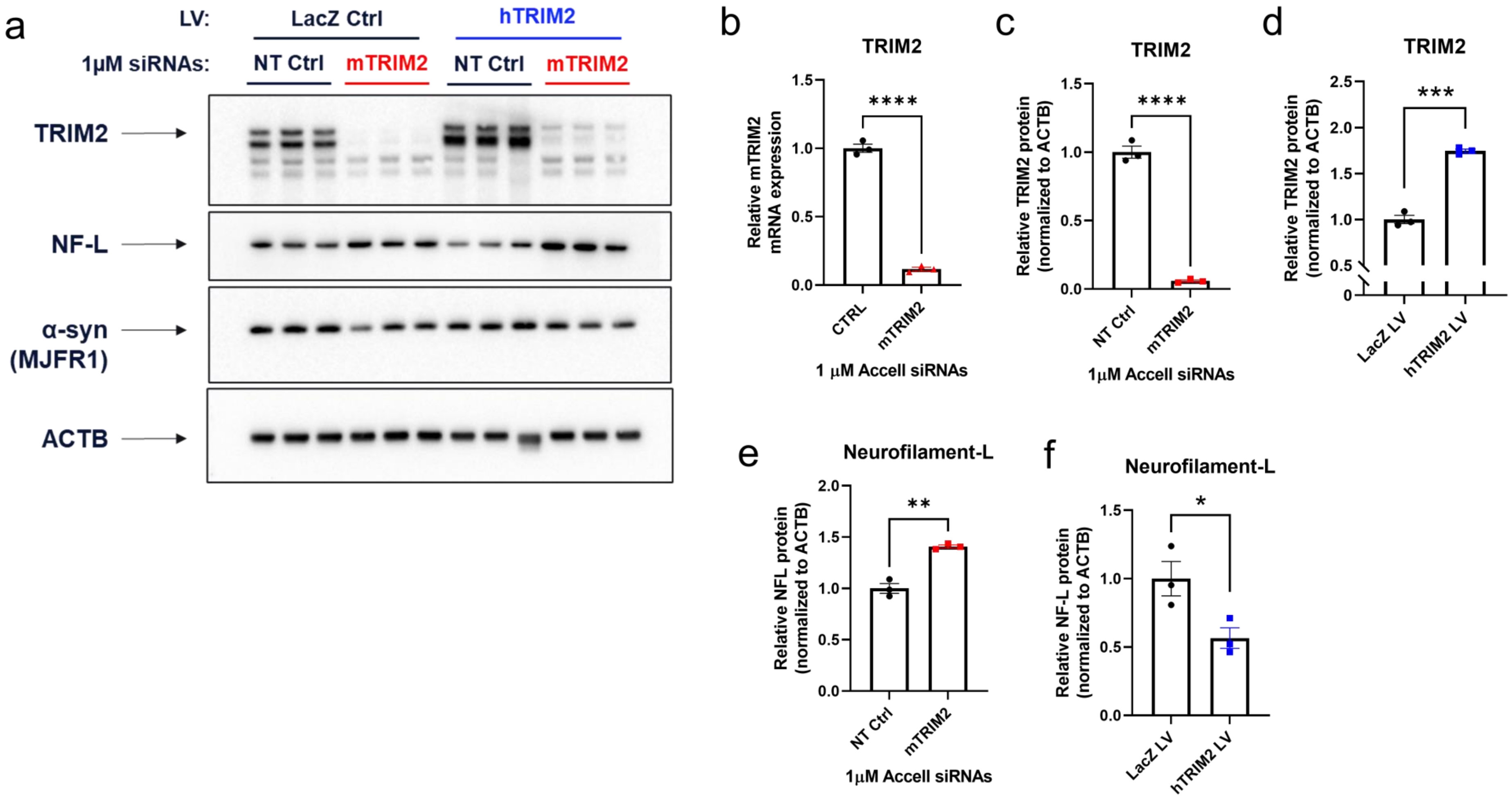

**Supplementary Fig 6. α-Syn preformed fibril treatment induces *de novo* aggregation of endogenous α-syn. a.** Representative images of 21 days in vitro (DIV) CD1 neurons treated with 1 µg/ml human α-syn preformed fibrils (PFFs) for 14 days and immunostained for DAPI (blue), NeuN (green), MAP2 (orange), and pS129 α-syn (red). Note the absence of pS129 α-syn immunostaining in control cells not treated with PFFs (left panels). **b.** Quantification of relative α-syn (SNCA) mRNA transcript levels in primary neurons treated with 1 µM Accell siRNAs targeting endogenous murine α-syn for 10 days, as determined by qPCR. ***$p<0.0001$ by unpaired Student's t-test; n=3/group. **c-d.** Quantification of PFF-induced pS129 α-syn pathology in CD1 (c) and M83 (d) neurons with and without Accell siRNA knockdown of endogenous murine (c) or human (d) α-syn. ***$p<0.0001$ by unpaired Student's t-test; n=6 wells/group. e. Liner regression analysis of NF-L intensity versus pS129 α-syn intensity per MAP2+ area within the same wells of M83 neurons treated with 1 µg/ml PFFs for 14 days. ***$p=0.0007$; R2=0.9191. Error bars represent mean ± SEM.

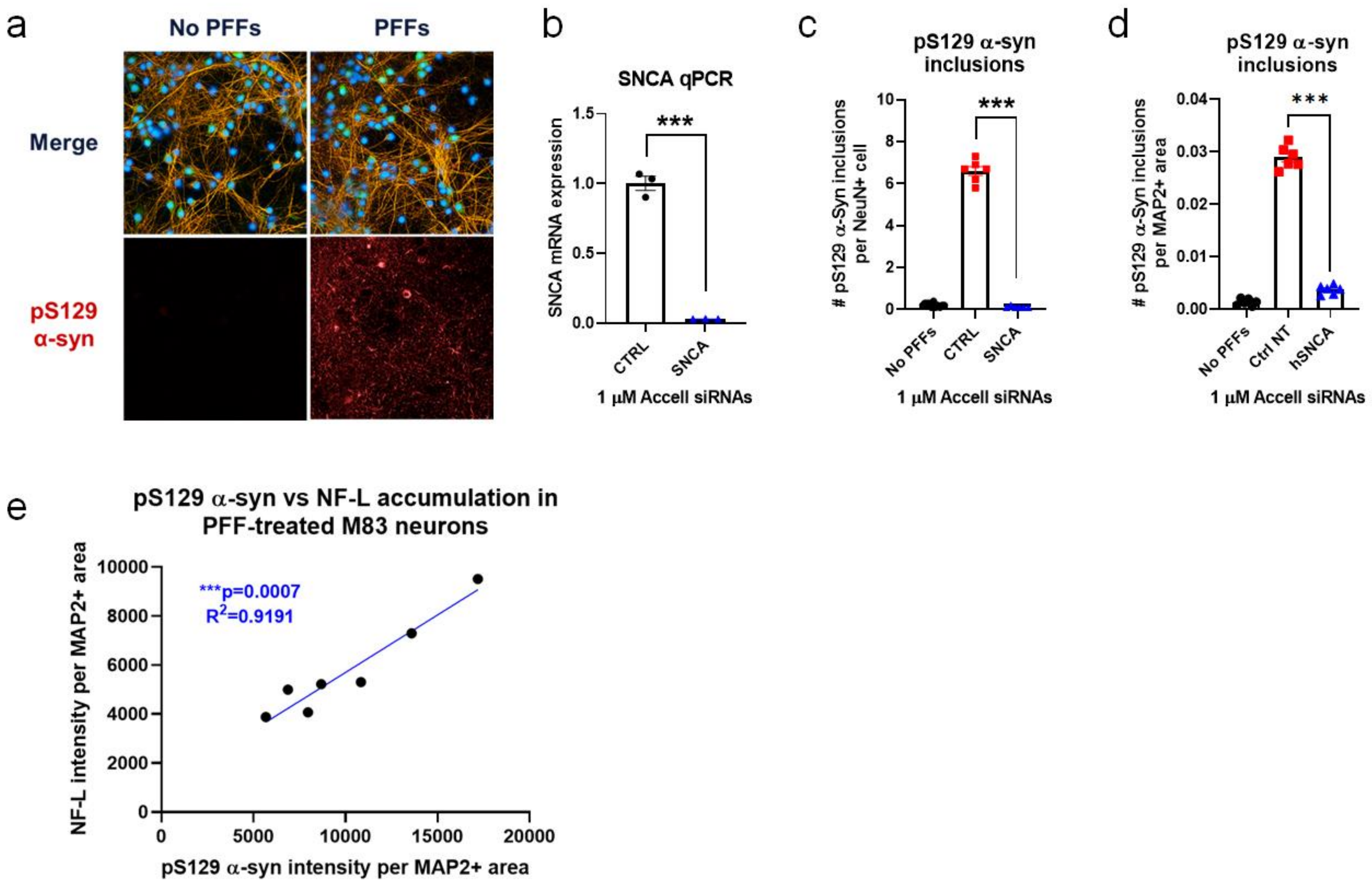

**Supplementary Fig 7. Time- and dose-dependent accumulation of pS129 α-syn pathology in preformed fibril-treated CD1 and M83 neurons. a-b**. Representative images of 21 days in vitro (DIV) CD1 (a, left panels) and M83 (b, right panels) neurons treated with various doses of human α-syn preformed fibrils (PFFs) for 7 or 14 days followed by immunostaining for DAPI (blue), NeuN (green), and pS129 α-syn (red). Images were captured at 40x magnification using a high-content confocal imaging system.

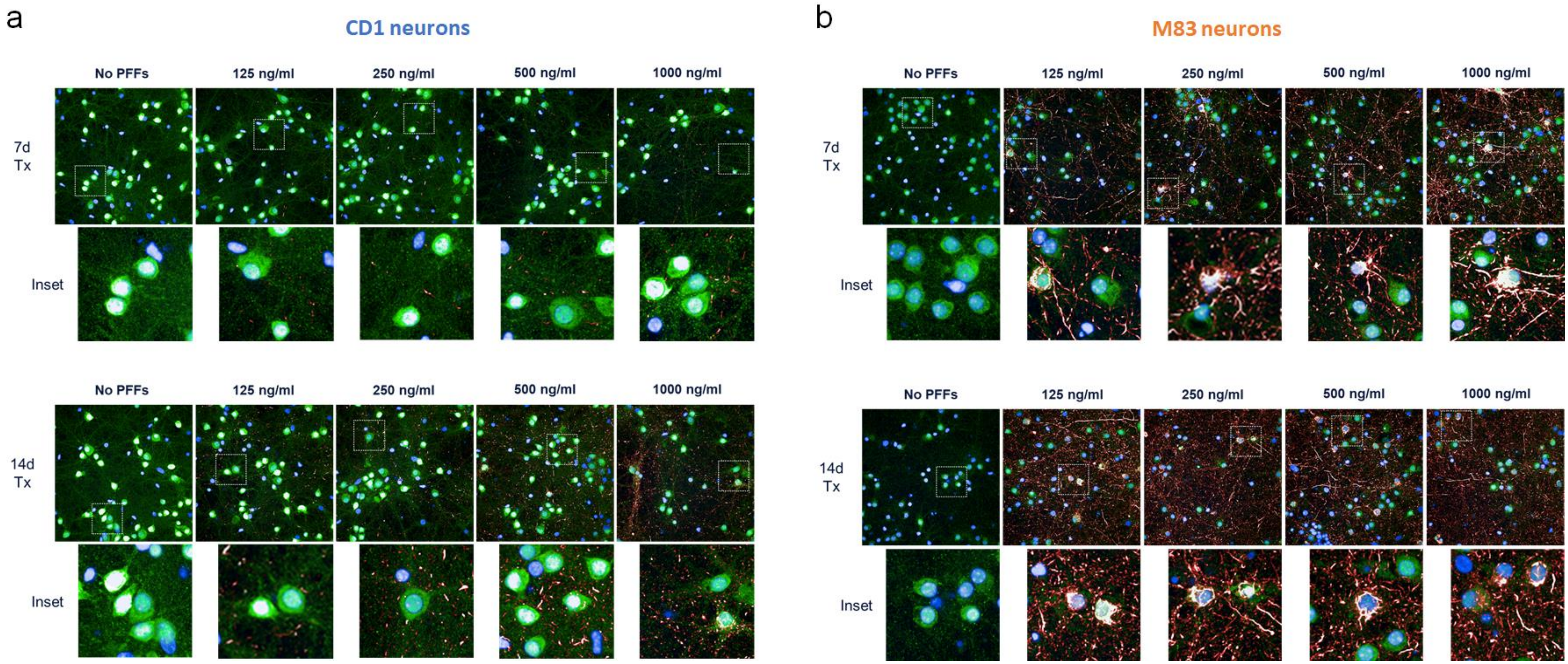

**Supplementary Fig 8. M83 neurons exhibit accelerated α-synuclein aggregation phenotype compared to CD1 neurons. a-h.** Quantification of total pS129 α-syn immunofluorescence intensity (a-b), number of pS129 α-syn inclusions (c-d), total pS129 α-syn aggregate area (e-f), and percentage of neurons (NeuN+ cells) with Lewy-like pS129 α-syn aggregates in the soma (g-h) in CD1 (left) and M83 (right) neurons treated with various concentrations of α-syn preformed fibrils (PFFs). Note the differences in the Y-axis between figures.

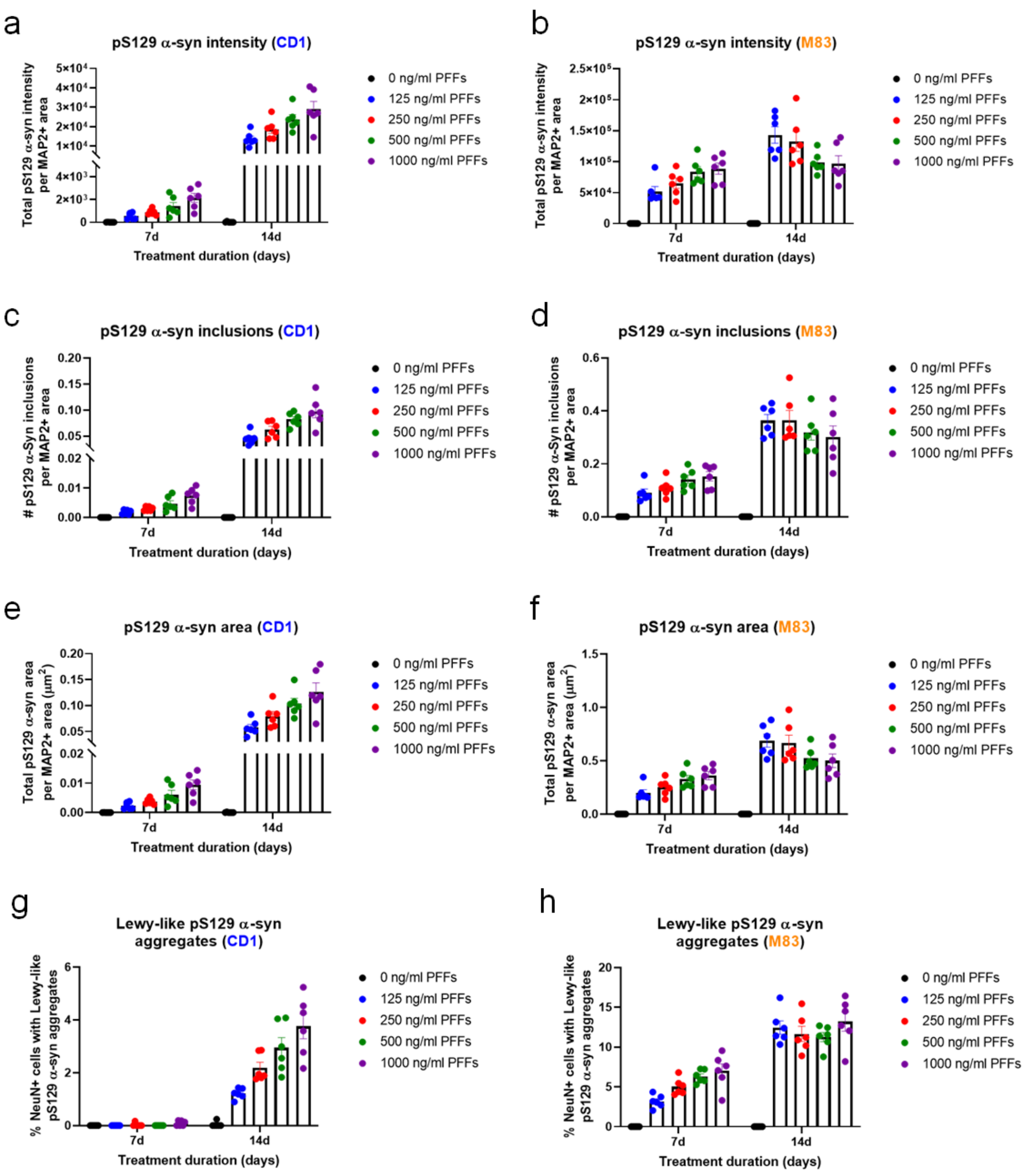

**Supplementary Fig 9. TRIM2 knockdown increases Lewy-like pS129 α-syn aggregates in M83 neurons.**
M83 mouse primary neurons were cultured for 21 days in vitro (DIV) with or without Accell siRNA knockdown of murine TRIM2 and treated with 250 ng/ml human α-syn preformed fibrils (PFFs) for the final 7 or 14 days. **a.** Quantification of the percentage of cells with Lewy-like pS129 α-syn aggregates in the neuronal cell body in M83 neurons treated with PFFs for 7 or 14 days. *p=0.0284 ***p=0.0010 by 2-Way ANOVA with Sidak's multiple comparisons test; n=15 wells/group. **b-c.** Quantification of the total pS129 α-syn immunofluorescence intensity (b) and area (c) for Lewy-like aggregates in the soma of M83 neurons treated with 1 µM control (CTRL) or TRIM2 targeting siRNAs followed by 250 ng/ml PFF treatment for 7d. **p=0.0020 (b) **p=0.0031 (c) by 2-Way ANOVA with Sidak's multiple comparison test; n=15 wells/group. **d-e.** Quantification of total MAP2+ neurite area per 40x field in M83 neurons treated with CTRL or TRIM2 targeting siRNAs ± 250 ng/ml PFFs for 7 (d) or 14 (e) days. **f-g.** Quantification of total pHrodo Red-conjugated PFF (pHrodo-PFF) intensity in M83 neurons treated with 1 µg/ml pHrodo-PFFs for 48h. pHrodo Red dye is non-fluorescent at neutral pH and exhibits increased fluorescence intensity only when trafficked into the acidic endo-lysosomal cellular compartment. Note that pHrodo-PFF uptake is not significantly different from control in cells treated with TRIM2 siRNAs (f) or transduced with hTRIM2 LV (g). Error bars represent mean ± SEM.

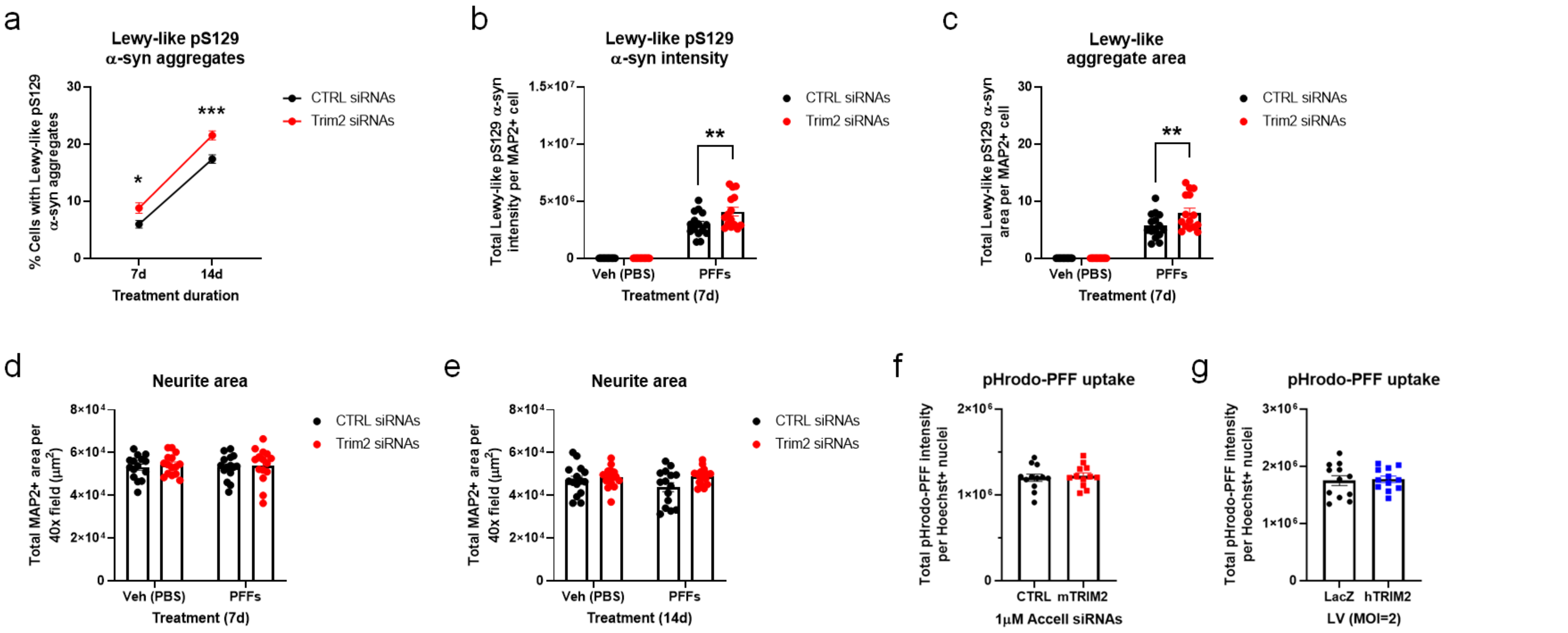

**Supplementary Fig 10. TRIM2 knockdown impairs lysosomal β-glucocerebrosidase enzymatic activity.**
**a.** Representative images of 18 days in vitro (DIV) M83 neurons incubated with 5 nM of the fluorescent β-glucocerebrosidase (GCase) activity-based probe MDW941 (orange) and Hoechst nuclear stain (blue) for 2h ± 1h pre-treatment with the irreversible GCase inhibitor CBE. Arrows denote cells magnified in the inset. Scale bar = 50µm. **b-c.** Quantification of total MDW941 fluorescence intensity per DAPI+ cell (b) and percentage of lysosomes containing active GCase molecules (c) in a. *p=0.0231 ***p=0.0003 by unpaired Student's t-test; n=5 wells/group. Error bars represent mean ± SEM.

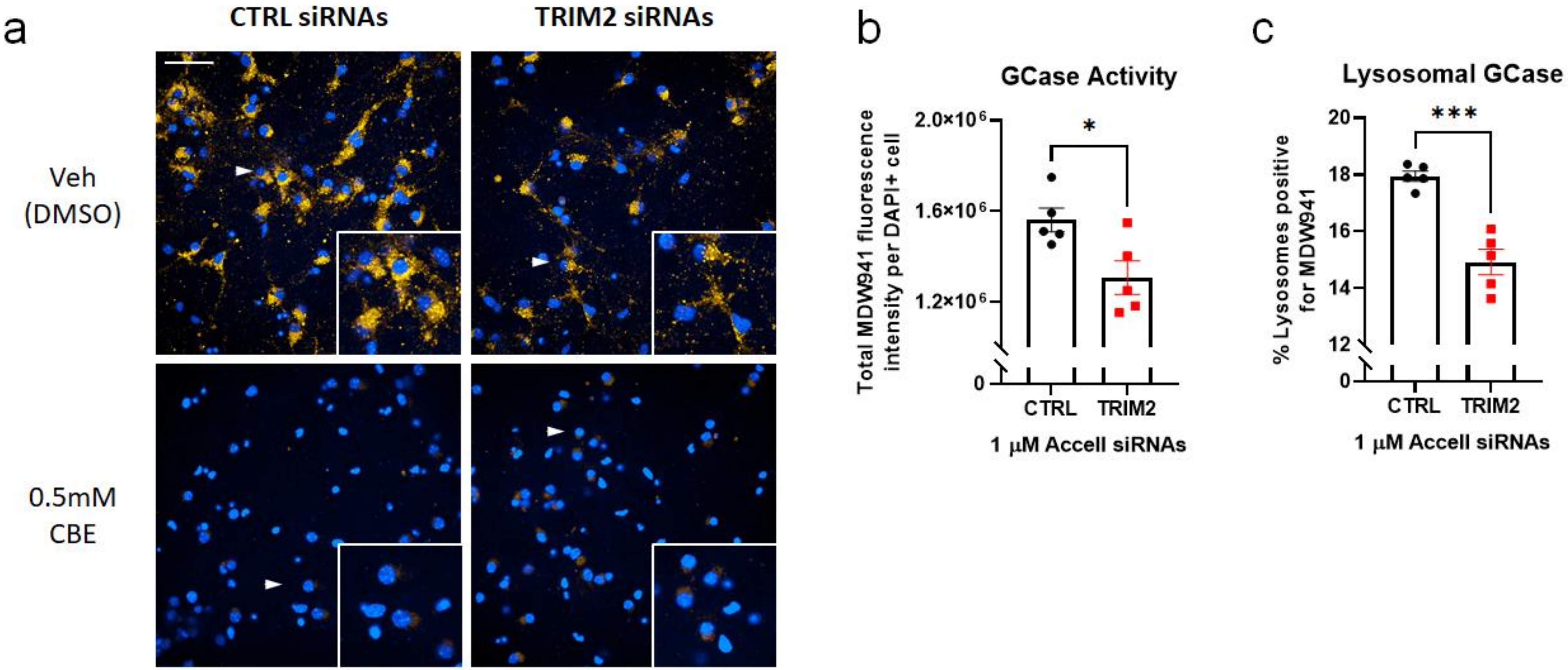

**Supplementary Fig 11. TRIM2 knockdown increases mitochondria number and decreases mitochondria size. a.** Representative images of 18 days in vitro (DIV) M83 neurons incubated with 50 nM MitoTracker Green dye (green) and Hoechst nuclear stain (blue) for 30 min. Images were processed using the SER ridge filtering algorithm in Perkin Elmer Harmony image analysis software to enable quantification of the morphological properties of the mitochondrial network (right panels). Scale bar = 20µm. **b-c.** Quantification of number (b) and size (c) of MitoTracker Green positive objects in a. **p=0.0032 (b) **p=0.0010 by unpaired Student's t-test; n=5 wells/group. **d.** Quantification of the percentage of mitochondria exhibiting fragmented versus elongated morphology in a. ***p=0.0005 by unpaired Student's t-test; n=5 wells/group. Error bars represent mean ± SEM.

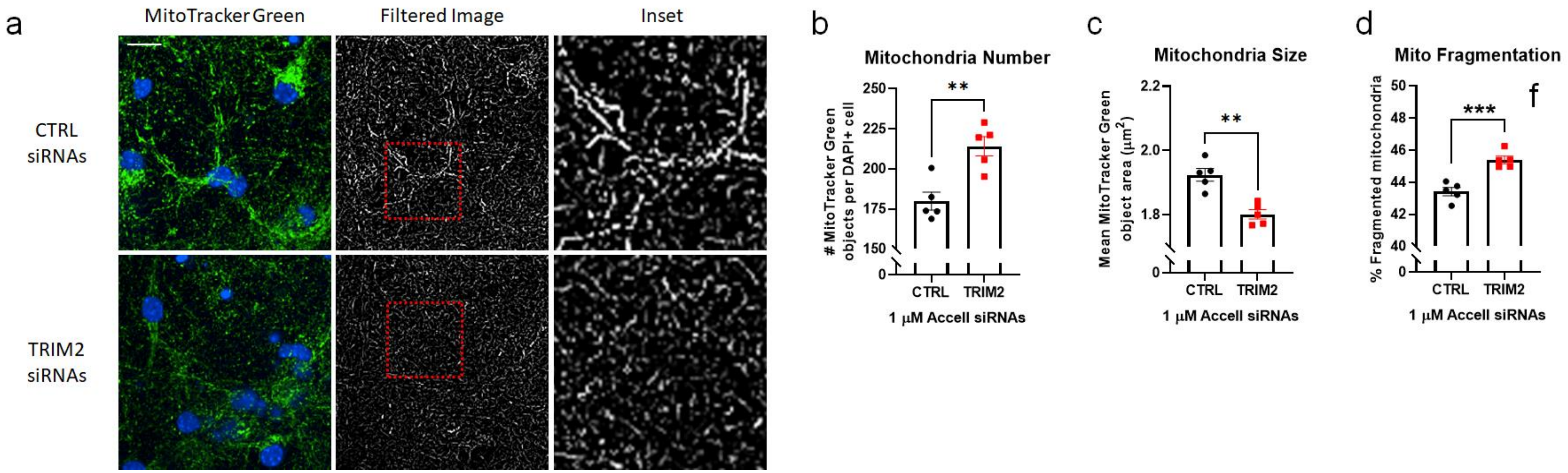

**Supplementary Fig 12. TRIM2 variant carriers show increased neurofilament-L protein level in plasma and CSF.** Comparisons of peripheral NF-L protein levels in PD patients from PPMI and PDBP cohorts with variants in TRIM2 identified from PPMI GWAS **a.** Plasma NF-L level stratified by rs72726947 alleles **b.** CSF NF-L level stratified by rs62323742 alleles. CSF NF-L level was significantly higher in GA than in GG (p=0.007, Welch's t-test). Note that statistical test could not be performed for AA group as there are only two data points. **c.** Plasma NF-L level stratified by rs62323742 alleles. CSF NF-L level was significantly higher in GG than in AA (p=0.06, Welch's t-test) **d.** CSF NF-L level stratified by rs62323742 alleles. Homozygous for reference allele (green), heterozygous (orange) and homozygous for alternative allele (purple).

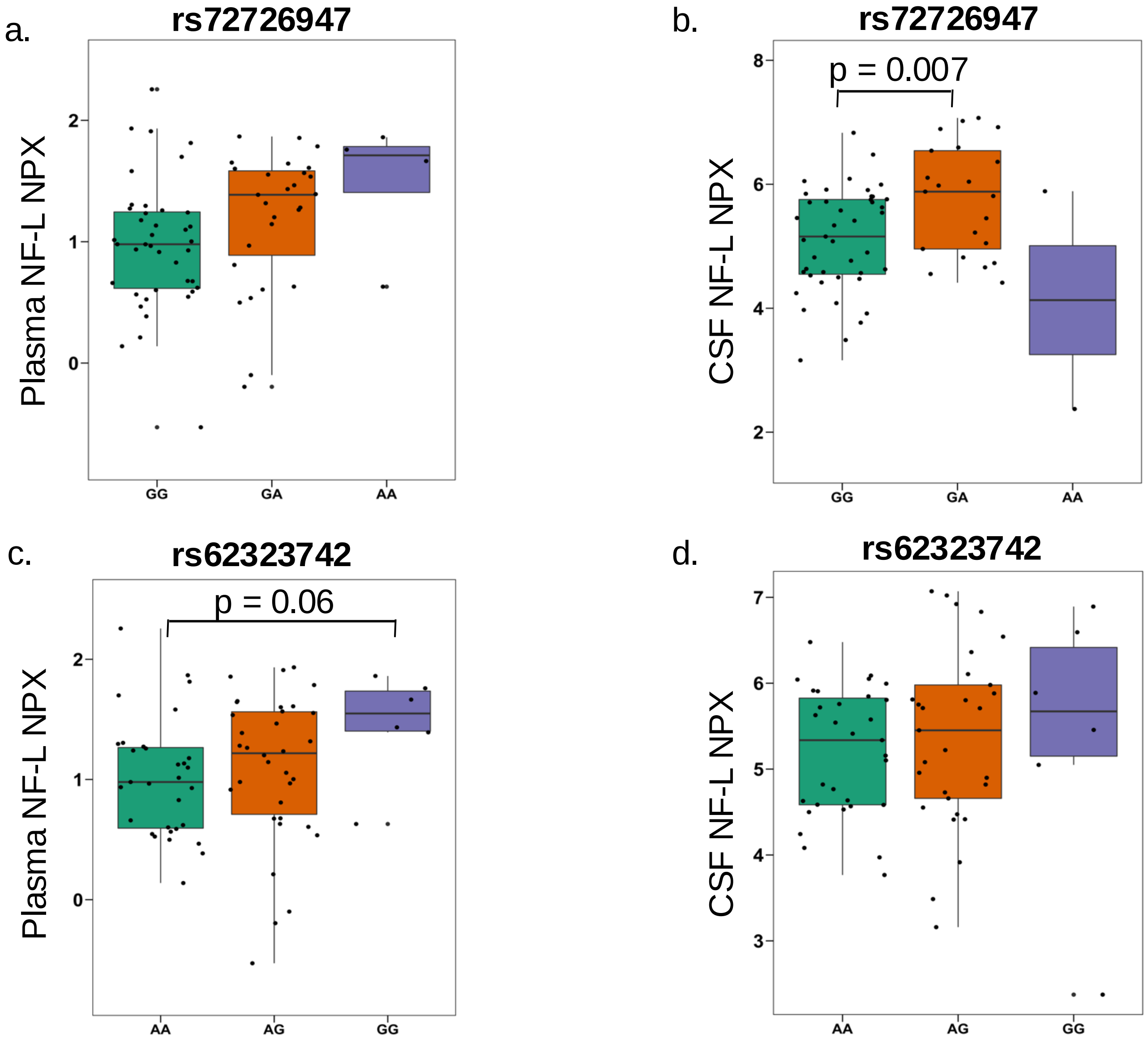
